# Mediating Effects of Healthy Lifestyle Factors on Associations between Mental Health and Functional Outcomes in Early Adolescence

**DOI:** 10.64898/2026.02.10.26345879

**Authors:** Jason Smucny, Tyler A. Lesh, Tara A. Niendam, Nicole R. Karcher

## Abstract

**Objective:** Although mental health and healthy lifestyle interventions are associated with functional outcomes in adolescence, the extent to which particular lifestyle factors explain relationships between mental health and outcome are unclear. Here we examined mediating effects of lifestyle factors on relationships between mental health and two functional outcomes measured 2-3 years later as well as the moderating effect of environmental risk factors on mediation strength in early adolescence.

**Methods:** This study analyzed data from 3 waves of the Adolescent Brain Cognitive Development Study (ages 10-11, 11-12, and 12-13). Mediating effects of sleep quality, screen time, physical activity and Mediterranean diet on the relationships between depression, anxiety, psychotic-like experience (PLE) distress, and total problems with two subsequent functional outcomes (academic functioning and social problems) were examined. Secondary analyses included environmental factors as moderators.

**Results:** Sleep quality mediated 18.5%, 36.3%, 8.3%, and 3.4% of the relationships between depression, anxiety, PLE distress and total problems with academic functioning, respectively. Screen time was the second strongest mediating factor. For social problems, only sleep quality showed > 3% mediation (19.6% – 23.3%). Mediating effects of sleep and screen time on academic functioning decreased as financial adversity increased. Conversely, mediating effects of sleep quality on social problems increased with worsening family conflict, financial adversity, and school environment.

**Conclusions:** These results suggest that healthy lifestyle factors (in particular sleep quality) may partially explain the associations between mental health and functioning in adolescents and suggest that these effects are modulated by environmental factors. These results may have important implications for future intervention studies.

## Introduction

Adolescence represents a critical developmental window during which mental health concerns, including depression, anxiety, and psychotic-like experiences (PLEs), often emerge (1). Globally, approximately one in seven 10-to 19-year-olds experience a mental disorder, and depression and anxiety are leading causes of illness and disability among young people (2). Understanding both the functional correlates of early mental health concerns and the targetable mechanisms underlying their development is of great importance for both public health and developmental science. The current paper directly addresses this need by examining potentially targetable mechanisms linked to the emergence of early mental health concerns.

Mental health difficulties experienced in adolescence often have a number of downstream consequences, including impact on everyday functioning such as academic achievement and peer/social functioning (3, 4). Mood and anxiety disorders have been associated with school absenteeism, lower grades, and withdrawal from peer relationships (5–9). Likewise, psychotic-like experiences (PLEs) may disrupt social engagement, peer support, and academic performance (10, 11). Failure to examine functioning alongside symptomatology risks overlooking the real-world consequences of adolescent mental health concerns and misses opportunities to intervene early and prevent these potential downstream impacts.

Given the relationship between early mental health concerns and everyday functioning, elucidating mechanisms underlying these associations becomes imperative. Among readily modifiable mechanisms, lifestyle factors, including healthy sleep, physical activity, diet quality, and screentime hold promise as leverageable targets in prevention and intervention. Research in “lifestyle psychiatry” has underscored that physical activity, sleep, and diet contribute meaningfully to the onset, maintenance, and alleviation of mental disorders (12). For example, higher levels of exercise have been linked to reduced depressive and anxiety symptoms in youth, and emerging evidence also points to benefits for social and academic functioning (13). Sleep disruptions, including but not limited to insufficient duration, poor quality, or irregular timing, have been associated with increased internalizing symptoms and poorer school and interpersonal functioning in adolescents (14, 15). Diet quality, especially adherence to a Mediterranean diet, has been connected to lower levels of depressive and anxiety symptoms in children and adolescents (16). Screen time presents another critical lifestyle parameter: extensive screen exposure, especially via social media and passive screen use, has been consistently tied to worse mental health outcomes in adolescents, and may further impair social and academic engagement through sleep displacement, reduced activity, and diminished in-person peer interaction (17, 18). Consistent with this, research using the Adolescent Brain Cognitive Development (ABCD) study has found evidence that increased screen time (19), greater sleep disturbances (20), and lower physical activity (21) are associated with greater endorsement of mental health concerns. The current manuscript aims to examine the untested model that these lifestyle behaviors act as mediators that link adolescent mental health (e.g., depression, anxiety, and PLEs) to everyday academic and social functioning.

It is also likely that these lifestyle mechanisms (i.e., sleep, physical activity, diet, and screen time) do not operate uniformly across all youth but rather are shaped by environmental context. For instance, environmental risk factors including financial adversity (e.g., socioeconomic strain), neighborhood and school conditions, and/or familial conflict may moderate the strength of the pathways linking lifestyle behaviors to mental health and functioning. Adolescents growing up in economically disadvantaged conditions (22, 23) or high-conflict homes (24) may face compounding risk, such that the same amount of sleep disruption or elevated screen time may confer greater detriment in the presence of environmental risk factors, or lifestyle buffers such as exercise may be undermined by environmental risk factors. The possibility of moderated mediation, whereby the environment (e.g., family conflict, disadvantage) influences the degree to which lifestyle factors mediate associations between symptoms with functioning, is an unexplored yet potentially compelling framework for understanding heterogeneity in adolescent risk for functioning decline.

In the present study, we leveraged data from the longitudinal, multi-site ABCD Study to test a conceptual model whereby adolescent depression, anxiety, and PLEs predict academic and social functioning, with lifestyle behaviors (screen time, sleep quality, physical activity, and diet) acting as mediators partially linking these associations. Although mediating effects of many interventions could be examined, we focused on these four lifestyle factors not only because of their established links to functioning but also because they can be addressed in outpatient therapeutic settings. Follow-up analyses explored the possibility of moderated mediation, exploring whether links between lifestyle factors and functioning were moderated by environmental risk factors (i.e., financial adversity, neighborhood safety, school quality, and familial conflict). This work aims to provide novel evidence identifying actionable targets for early intervention and to delineate under which conditions healthy lifestyle factors are most impactful.

## Methods

### Participants

ABCD data used in this report came from release 5.1 (DOI: 10.15154/z563-zd24). DOIs can be found at https://nda.nih.gov/abcd/abcd-annual-releases.html. The present study primarily used 1-year follow-up data (participant ages 10-11) as clinical predictors, 2-year follow-up data (ages 11-12) as mediators (except for Mediterranean diet, which was only available at 1-year follow-up), and 3-year follow-up data (ages 12-13) as the outcome measure in mediation models. Exclusion criteria are provided in the Supplement.

### Mental Health Predictors

The following mental health measures were included as predictors in mediation models: Child Behavior Checklist (CBCL) (3) Depression T-score, CBCL Anxiety T-score, CBCL Total Problems T-score (a composite of all CBCL ratings), and Prodromal Questionnaire-Brief Child Version (PQ-BC) PLEs distress score sum (4). Depression and anxiety scores were taken from the DSM-5-oriented affective problems and anxiety problems CBCL scales. All predictive clinical measures were taken from 1-year follow-up data.

### Healthy Lifestyle Mediators

The following “healthy lifestyle” measures were included as potential mediators in analyses: diet, physical activity, screen time, and sleep quality. “Diet” was captured by the parent-reported sum of the Mediterranean-DASH Intervention for Neurodegenerative Delay (MIND) diet questionnaire (see Supplement) (15). “Physical activity” was captured by the child’s answer to the question: “During the past 7 days, on how many days were you physically active for a total of at least 60 minutes per day?” (from 0 to 7). “Screen time” was captured by parent-reported screen time on weekends or weekdays. Weekend and weekday screen time were examined separately because they may reflect different aspects of time allocation (i.e., recreational time allocation vs. educational (“occupational”) time allocation). “Sleep quality” was captured by taking the caregiver-reported sum of the Sleep Disturbance Scale for Children (SDSC) (14) (see Supplement). Screen time and sleep quality measures were reverse coded such that greater values on all scales corresponded to greater adherence towards following healthy lifestyle habits. Physical activity, screen time, and sleep quality were taken from 2-year follow-up data. Diet information was taken from 1-year follow-up data (as it was not included as part of 2-year follow-up data; notably, year-to-year diets are unlikely to substantially change between ages 10-12 (25)).

### Functional Outcomes

Two functional outcomes were examined: academic functioning during follow-up years 2-3, and social problems at 3-year follow-up (i.e., between ages 12-13). Parents provided academic functioning inforatmion at 3-year follow-up by reporting their child’s average grades during the preceding year converted to a 1-12 scale, with 1 corresponding to “A+” or 97-100%, 2 with “A” or 93-97%, 3 with “A-” or 90-92%, etc. down to 12 with “F” or < 65%. These scores were reverse coded such that higher scores were associated with higher grades. Children whose parents refused to report grades were excluded from any analyses with grades. Social problems were quantified by CBCL Social Problems T-score, which includes questions regarding getting along with other children, getting teased, feeling lonely, and acting immaturely (26).

### Environmental Adversity Moderators

The following environmental adversity moderators were examined: neighborhood safety, family conflict, school environment, and family financial adversity. Neighborhood safety was captured by the parental report ABCD Neighborhood Crime and Safety Survey (27) (lower scores = safer neighborhood), family conflict by the ABCD Parent Family Environment Scale-Family Conflict Subscale (modified from PhenX) (28) (higher scores = more conflict), school environment by the youth report School Risk and Protective Factors School Environment subscale (modified from PhenX) (29) (higher scores = better environment), and family financial adversity by the summed endorsement of 7 parent-reported questions of financial difficulties experienced during the past 12 months from a demographic questionnaire (30) (higher scores = more adversity). More detailed information on these instruments is provided in the Supplement.

### Mediation Analyses

Mediation analyses were performed using the PROCESS v.5.0 toolbox (processmacro.org) in SPSS v.30 (IBM). The primary analyses included eight mediation models; each model included one of the mental health measures (e.g., depression) as the independent variable, age/sex/site as covariates, the four healthy lifestyle factors (e.g., sleep quality) as simultaneous (parallel) mediators, and one of two functional outcome measures (academic functioning and social problems) as the dependent variable. Total problems score was not included in models with social problems as the outcome due to overlap with social problems. Secondary analyses were performed similarly except they included NIH Cognition Toolbox Composite (31) raw score (an IQ measure) as a covariate as previous research indicates grades (32) and social functioning (33) may be influenced by IQ. A third set of analyses examined weekday screen time instead of weekend screen time. Independent variables were taken from 1-year follow-up data, mediators from 2-year follow-up (except for diet, which was only available at 1-year follow-up), and outcome measures from 3-year follow-up questionnaires. Statistical details are provided in the Supplement.

### Moderated Mediation Effects

Briefly, moderated mediation models examined the degree to which each moderator (e.g., family financial adversity) affected the mediation of associations between mental health with outcome by affecting the relationships between lifestyle factors and outcome. A diagram of the analysis strategy for environmental moderation effects is shown in Supplementary Figure 1. Additional information is provided in the Supplement.

## Results

### Sample

Demographic, clinical, behavioral, environmental, and functional information for children included in the study are presented in Table 1. Briefly, the analyses with academic functioning as the outcome included 6754 participants (after excluding siblings and children with missing data) and the analyses with social problems as the outcome included 7550 participants.

**Table 1.** Demographic, clinical, behavioral, environmental, and functional information for all participants included in analyses. Values are from parental report data unless otherwise specified (parental report data was used unless unavailable). Numbers in parentheses represent the standard deviation. *A grades score of 3 corresponds to a GPA of 3.5-3.69 (A-) and a score of 4 to a GPA of 3.30-3.49 (B+). CBCL = Child Behavior Checklist, NIH = National Institutes of Health, PQ-BC = Prodromal Questionnaire-Brief Child Version.

| Analysis Dependent Variable | Grades in School, Years 2-3 | CBCL Social Problems Score, Year 3 |
| --- | --- | --- |
| **N** | 6754 | 7550 |
| **Demographic Covariates** |  |  |
| Age in Years at Year 2 | 11.99 (.66) | 12.00 (.66) |
| Sex *n* M/F/Intersex | 3578/3174/2 | 3988/3560/2 |
| NIH Cognition Toolbox Total Composite Raw Score, Baseline | 87.11 (8.89) | 86.79 (9.01) |
| **Clinical, Independent (Predictor) Variables** |  |  |
| CBCL Depression T-Score, Year 1 | 53.87 (5.97) | 53.97 (6.08) |
| CBCL Anxiety T-Score, Year 1 | 53.55 (6.10) | 53.63 (6.25) |
| PQ-BC Distress Score, Year 1 | 4.40 (9.05) | 4.48 (9.09) |
| CBCL Total Problems T-Score, Year 1 | 45.51 (10.95) | 45.65 (11.14) |
| **Healthy Lifestyle Mediators** |  |  |
| Sleep Disturbances Score, Year 2 | 36.43 (8.01) | 36.48 (8.06) |
| Mediterranean Diet Score, Year 1 | 8.08 (2.39) | 8.07 (2.42) |
| Youth Reported Days Physically Active in the Last Week, Year 2 Questionnaire | 3.80 (2.15) | 3.78 (2.16) |
| Weekend Screen Time Hours, Year 2 | 4.63 (3.07) | 4.69 (3.13) |
| Weekday Screen Time Hours, Year 2 | 3.30 (3.08) | 3.38 (3.15) |
| **Environmental Moderators** |  |  |
| Neighborhood Crime Rating, Year 2 | 3.88 (.93) | 3.87 (.94) |
| Family Conflict Score, Year 2 | 2.36 (1.94) | 2.38 (1.96) |
| Youth Reported School Environment Score, Year 2 | 19.63 (2.76) | 19.63 (2.78) |
| Family Financial Adversity Score, Baseline | .40 (1.01) | .42 (1.05) |
| **Functional, Dependent (Outcome) Variables** |  |  |
| Grades in School*, Years 2-3 | 3.73 (2.31) | — |
| CBCL Social Problems Score, Year 3 | — | 52.65 (4.90) |

### Mediation Analyses: Mental Health Predicting Academic Functioning

First, we ran a set of mediation analyses in which a mental health measure (depression, anxiety, distressing PLEs, and CBCL total problems) was the independent variable, four healthy lifestyle factors (sleep quality, weekend screen time, diet, and physical activity) were mediators, sex/age/site were covariates, and academic functioning (school grades) was the outcome. When entered simultaneously, these lifestyle factors combined significantly mediated associations between all four mental health variables with academic functioning (see Figure 1 for mediation models and Supplementary Table 1 for detailed statistics). The relationship between anxiety and academic functioning showed the highest mediation from these lifestyle mediators (48%), followed by depression (30%), distressing PLEs (18%), and finally total problems (10%). Furthermore, in all four models, sleep showed evidence of being the most important mediator, showing 36.3% (of 48% total) mediation for anxiety, 18.5% (of 30% total) mediation for depression, 8.3% (of 18% total) mediation for distressing PLEs, and 4.6% (of 10% total) mediation for total problems (Figure 1, Supplementary Table 1). Screen time was the second-most important mediator, although its contributions were often noticeably less than sleep quality (e.g., 36.3% and 18.5% mediation by sleep quality vs. 5.0% and 6.3% mediation by screen time when anxiety and depression were the independent variables, respectively). Exercise also showed significant mediation, although its effects were even smaller than for screen time (e.g., 2.8% and 4.0% mediation for models predicting anxiety and depression, respectively). Diet did not show significant mediation for depression, distressing PLEs, or total problems.

**Figure 1.**
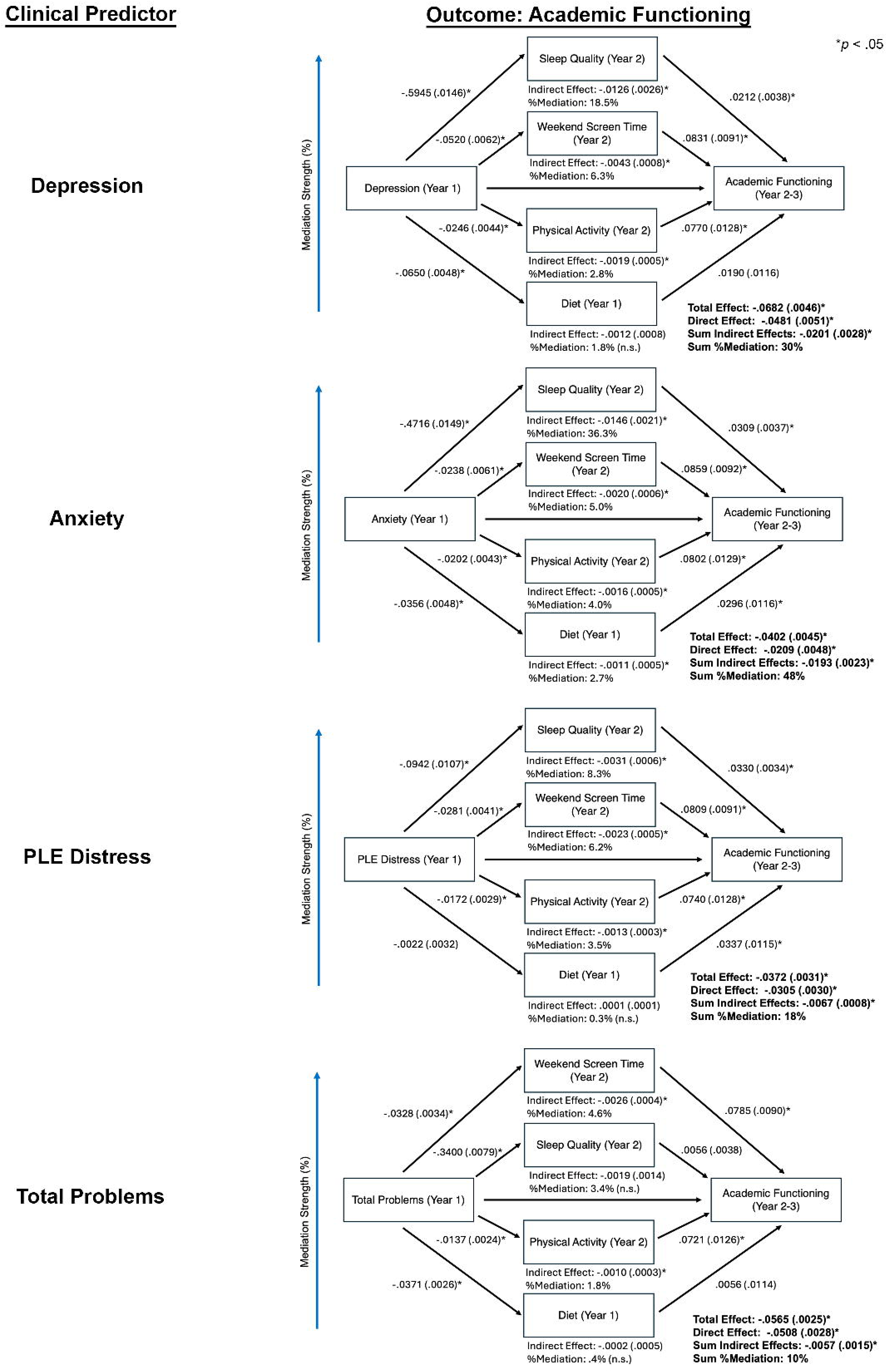
Results of mediation models predicting academic functioning (school grades between 2-3 year follow-up) while including age (2-year follow-up), sex, and site as covariates. More detailed results are provided in Supplementary Table 1. PLE = Psychotic-like Experiences.

Results from analyses that included NIH Cognition Toolbox Composite score as a covariate and weekday instead of weekend screen time as a mediator are provided in the Supplement. Briefly, including cognition as a covariate did not appreciably alter the results, and mediating effects of weekly screen time were smaller but still statistically significant.

### Mediation Analyses: Mental Health Predicting Social Functioning

Next, we ran a set of mediation analyses in which a mental health measure was the independent variable, the four healthy lifestyle factors were mediators, sex/age/site were covariates, and social problems (CBCL Social Problems score) was the outcome. As was the case for academic functioning, the four lifestyle mediators combined to show significant mediation in all psychopathological models (depression, anxiety, and distressing PLEs; Figure 2, Supplementary Table 4). The relationship between distressing PLEs and social problems showed the highest mediation from these lifestyle mediators (27%), followed by depression (21%) and anxiety (21%). Strikingly, amongst the lifestyle mediators, only sleep quality was a robust, consistently significant mediator for all models examined. More specifically, for depression, sleep showed 19.9% (of 21% total) mediation, for anxiety sleep showed 19.6% (of 21% total) mediation, and for PLE distress sleep showed 23.3% (of 27% total) mediation. Thus, almost all of the healthy lifestyle mediation effects were driven by sleep quality for all three mental health domains examined.

**Figure 2.**
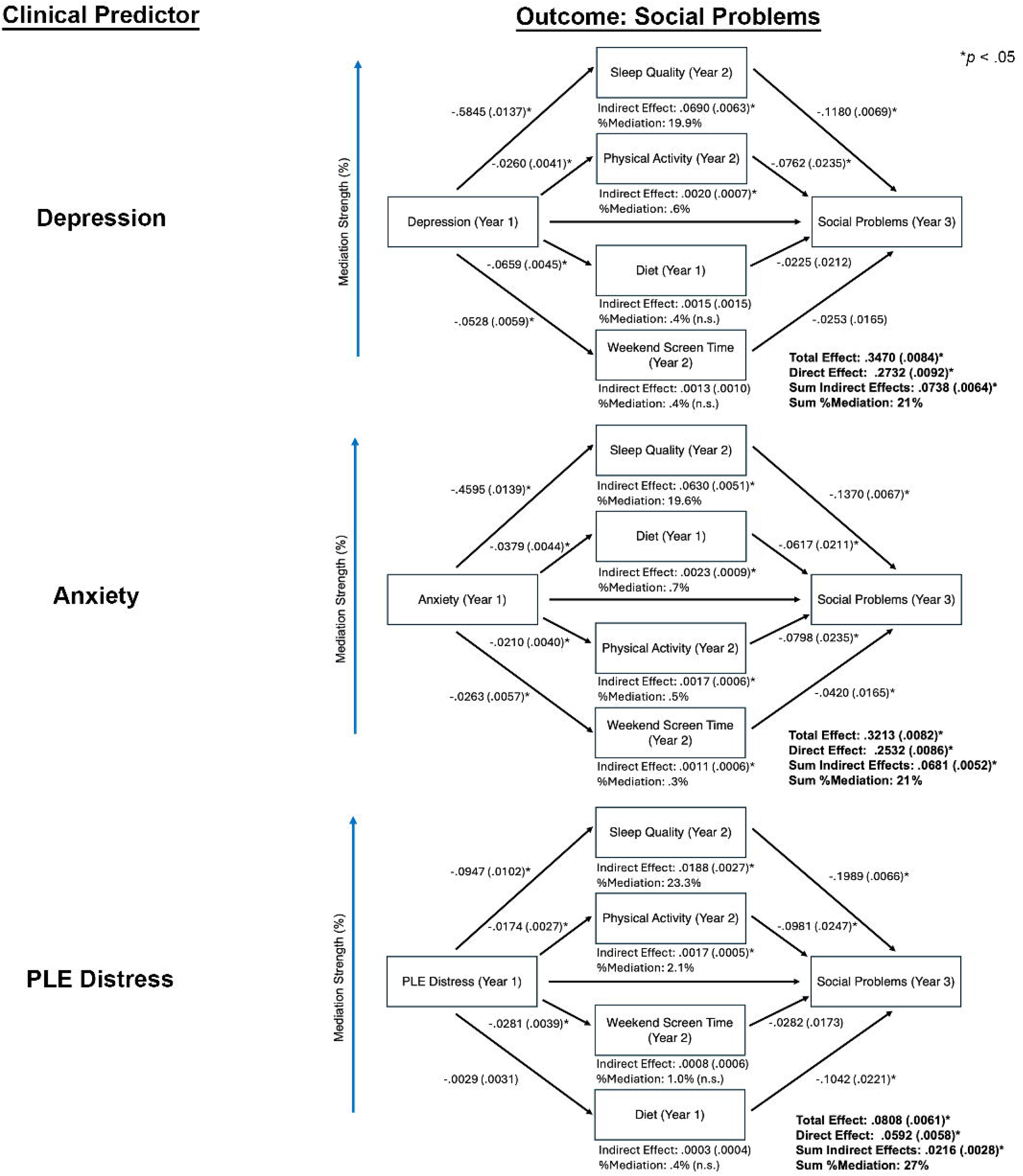
Results of mediation models predicting social problems (3 year CBCL Social Problems score) while including age (2-year follow-up), sex, and site as covariates. More detailed results are provided in Supplementary Table 4. PLE = Psychotic-like Experiences.

Results using cognition as a covariate and weekday instead of weekend screen time as a mediator are provided in the Supplement. Briefly, including cognition as a covariate did not appreciably alter the results, and mediating effects of weekly screen time became smaller and statistically non-significant.

### Environmental Moderation Effects

Supplementary Table 7 depicts how environmental factors moderate associations between lifestyle mediators with academic functioning, as well as moderator effects on percent mediation. As can be seen in Table 2, as financial adversity increased, the mediating role of sleep quality and/or weekend screen time (reverse-scored) on associations between mental health and academic functioning decreased.

**Table 2.**
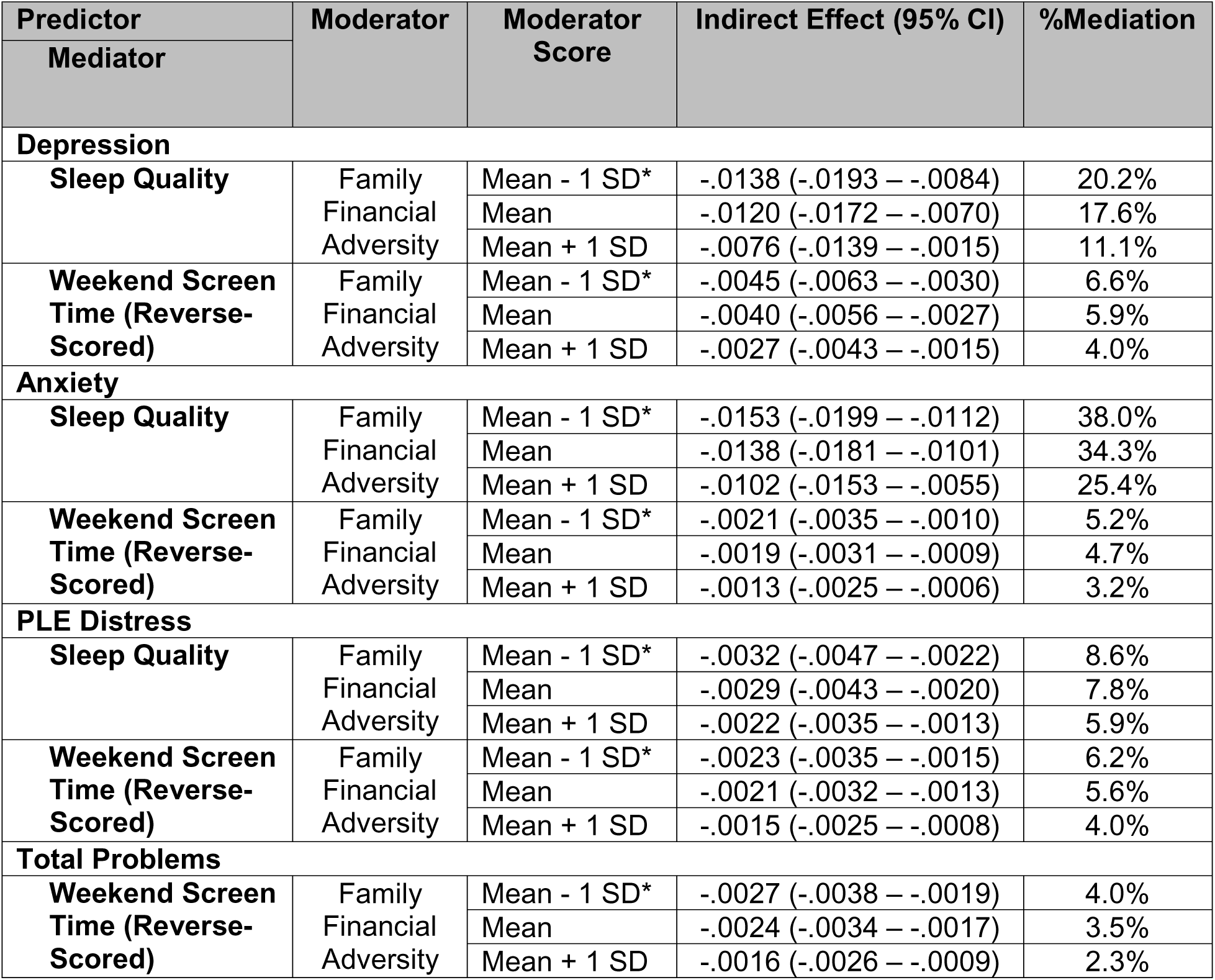
Changes in %mediation dependent on moderator level for significant moderators for models predicting academic functioning between years 2-3. See Table 1 for mean (SD) of moderator variables. *Because mean - 1 SD < 0, mediation effects at this level were calculated when the moderator = 0.

| Predictor<br>Mediator | Moderator | Moderator<br>Score | Indirect Effect (95% CI) | %Mediation |
| --- | --- | --- | --- | --- |
| Depression |  |  |  |  |
| Sleep Quality | Family<br>Financial<br>Adversity | Mean - 1 SD* | -.0138 (-.0193 – -.0084) | 20.2% |
|  |  | Mean | -.0120 (-.0172 – -.0070) | 17.6% |
|  |  | Mean + 1 SD | -.0076 (-.0139 – -.0015) | 11.1% |
| Weekend Screen<br>Time (Reverse-Scored) | Family<br>Financial<br>Adversity | Mean - 1 SD* | -.0045 (-.0063 – -.0030) | 6.6% |
|  |  | Mean | -.0040 (-.0056 – -.0027) | 5.9% |
|  |  | Mean + 1 SD | -.0027 (-.0043 – -.0015) | 4.0% |
| Anxiety |  |  |  |  |
| Sleep Quality | Family<br>Financial<br>Adversity | Mean - 1 SD* | -.0153 (-.0199 – -.0112) | 38.0% |
|  |  | Mean | -.0138 (-.0181 – -.0101) | 34.3% |
|  |  | Mean + 1 SD | -.0102 (-.0153 – -.0055) | 25.4% |
| Weekend Screen<br>Time (Reverse-Scored) | Family<br>Financial<br>Adversity | Mean - 1 SD* | -.0021 (-.0035 – -.0010) | 5.2% |
|  |  | Mean | -.0019 (-.0031 – -.0009) | 4.7% |
|  |  | Mean + 1 SD | -.0013 (-.0025 – -.0006) | 3.2% |
| PLE Distress |  |  |  |  |
| Sleep Quality | Family<br>Financial<br>Adversity | Mean - 1 SD* | -.0032 (-.0047 – -.0022) | 8.6% |
|  |  | Mean | -.0029 (-.0043 – -.0020) | 7.8% |
|  |  | Mean + 1 SD | -.0022 (-.0035 – -.0013) | 5.9% |
| Weekend Screen<br>Time (Reverse-Scored) | Family<br>Financial<br>Adversity | Mean - 1 SD* | -.0023 (-.0035 – -.0015) | 6.2% |
|  |  | Mean | -.0021 (-.0032 – -.0013) | 5.6% |
|  |  | Mean + 1 SD | -.0015 (-.0025 – -.0008) | 4.0% |
| Total Problems |  |  |  |  |
| Weekend Screen<br>Time (Reverse-Scored) | Family<br>Financial<br>Adversity | Mean - 1 SD* | -.0027 (-.0038 – -.0019) | 4.0% |
|  |  | Mean | -.0024 (-.0034 – -.0017) | 3.5% |
|  |  | Mean + 1 SD | -.0016 (-.0026 – -.0009) | 2.3% |

Supplementary Table 8 depicts how environmental factors moderate associations between lifestyle mediators with social problems was the outcome. As can be seen in Table 3, as family conflict and financial adversity increased and school environment worsened, the mediating effects of sleep quality on associations between mental health and social problems increased.

**Table 3.** Changes in %mediation dependent on moderator level for significant moderators for models predicting social problems (CBCL Social Problems Score) at year 3. See Table 1 for mean (SD) of moderator variables. *Because mean - 1 SD < 0, mediation effects at this level were calculated when the moderator = 0.

| Predictor<br>Mediator | Moderator | Moderator<br>Score | Indirect Effect B (95%<br>CI) | %Mediation |
| --- | --- | --- | --- | --- |
| Depression |  |  |  |  |
| Sleep Quality | Family<br>Conflict | Mean - 1 SD | .0455 (.0318 – .0603) | 13.1% |
|  |  | Mean | .0595 (.0484 – .0716) | 17.1% |
|  |  | Mean + 1 SD | .0735 (.0593 – .0890) | 21.2% |
| Sleep Quality | School<br>Environment | Mean - 1 SD | .0777 (.0632 – .0933) | 22.4% |
|  |  | Mean | .0667 (.0549 – .0795) | 19.2% |
|  |  | Mean + 1 SD | .0557 (.0408 – .0719) | 16.1% |
| Sleep Quality | Family<br>Financial<br>Adversity | Mean - 1 SD* | .0590 (.0463 – .0717) | 17.0% |
|  |  | Mean | .0639 (.0523 – .0761) | 18.4% |
|  |  | Mean + 1 SD | .0762 (.0617 – .0928) | 22.0% |
| Anxiety |  |  |  |  |
| Sleep Quality | Family<br>Conflict | Mean - 1 SD | .0443 (.0332 – .0554) | 13.8% |
|  |  | Mean | .0547 (.0457 – .0648) | 17.0% |
|  |  | Mean + 1 SD | .0651 (.0539 – .0777) | 20.3% |
| Sleep Quality | School<br>Environment | Mean - 1 SD | .0690 (.0576 – .0816) | 21.5% |
|  |  | Mean | .0613 (.0519 – .0719) | 19.1% |
|  |  | Mean + 1 SD | .0536 (.0419 – .0673) | 16.7% |
| Sleep Quality | Family<br>Financial<br>Adversity | Mean - 1 SD* | .0543 (.0451 – .0650) | 16.9% |
|  |  | Mean | .0584 (.0494 – .0685) | 18.2% |
|  |  | Mean + 1 SD | .0683 (.0564 – .0821) | 21.3% |
| PLE Distress |  |  |  |  |
| Sleep Quality | Family<br>Conflict | Mean - 1 SD* | .0139 (.0010 – .0188) | 17.2% |
|  |  | Mean | .0165 (.0121 – .0215) | 20.4% |
|  |  | Mean + 1 SD | .0191 (.0140 – .0251) | 23.6% |
| Sleep Quality | School<br>Environment | Mean - 1 SD* | .0202 (.0151 – .0265) | 25.0% |
|  |  | Mean | .0184 (.0137 – .0240) | 22.8% |
|  |  | Mean + 1 SD | .0167 (.0120 – .0223) | 20.7% |
| Sleep Quality | Family<br>Financial<br>Adversity | Mean - 1 SD* | .0172 (.0126 – .0224) | 21.3% |
|  |  | Mean | .0179 (.0133 – .0233) | 22.2% |
|  |  | Mean + 1 SD | .0196 (.0146 – .0257) | 24.3% |

## Discussion

The present study examined the degree to which potentially modifiable lifestyle behaviors, including sleep quality, physical activity, diet quality, and screen time, serve as mechanisms linking adolescent mental health symptoms to academic functioning and social problems, as well as whether these associations were moderated in the context of environmental adversity. Using data from the ABCD Study, we found evidence that healthy lifestyle factors, especially sleep, mediated associations between depression, anxiety, and PLEs with both academic and social functioning. We also found evidence that financial adversity modified associations between lifestyle factors with both academic and social problems, and that school environment and family conflict additionally modified associations with social problems. These findings clarify modifiable pathways that may contribute to functional decline in adolescence and identify contextual conditions under which lifestyle-focused interventions may be less effective.

### Lifestyle Mediators Linking Symptoms to Academic Functioning

Consistent with predictions, healthy lifestyle factors jointly mediated the associations between symptoms and academic functioning across all mental health domains examined. For academic functioning, mediation was strongest for models using anxiety as the predictor (48% of the total effect), followed by depression, PLEs, and total mental health concerns. This pattern may suggest that academic functioning is particularly sensitive to internalizing processes (e.g., cognitive rumination, worrying), domains that may be especially intertwined with lifestyle behaviors (6, 34). Sleep quality emerged as the dominant mediator across all analyses (i.e., explaining the majority of the total mediated effect), with screen time providing additional contributions. These findings point to sleep quality as a key mechanism through which early symptoms translate into reduced academic performance, consistent with evidence linking sleep irregularities to daytime fatigue, attention problems, and poorer executive functioning (6, 8, 34). Additionally, screen time showed a moderate influence while exercise and diet showed less robust mediation effects, with their small but significant contributions in some models. This highlights the value of a multi-behavioral approach to supporting academic outcomes (12, 13, 16). Notably, weekend screen time showed a qualitatively stronger mediating effect than weekend screen time. One possible reason for the discrepancy is the dynamic range for weekday screen time is lower (as children are in school, and as evidenced by the lower mean weekday screen time (Table 1)), reducing its ability to mediate functional outcomes. Overall, these results suggest that interventions that improve sleep quality and/or (to a lesser extent) restrict screen time may reduce the deleterious effects of mental health problems on academic functioning.

### Lifestyle Mediators Linking Symptoms to Social Problems

When social problems were the outcome, a similar overall mediation pattern emerged as compared to academic functioning, although the strength of the mediation differed. Mediation was strongest for PLE distress (27%), consistent with some psychosis spectrum research (10, 11), followed by depression and anxiety (both ∼21%) (5, 9). Unlike the academic functioning models where multiple healthy lifestyle factors contributed, sleep quality was the only consistently significant mediator of social functioning across all domains. This may indicate that sleep plays a uniquely central role in how symptoms translate into impairments in social functioning. This is potentially consistent with previous research linking mental health concerns, including social anhedonia and irritability, with social problems (35, 36). In contrast, screen time, diet, and physical activity contributed minimally to social problems pathways, suggesting these behaviors exert a smaller influence on associations between mental health with social problems. The contrast between academic functioning and social problems mediation patterns underscores that healthy lifestyle factors may operate differently across functional domains, with sleep exerting a more domain-general influence, and other lifestyle factors (e.g., screentime) playing more domain-specific roles. As with academic functioning, these results also suggest that interventions that improve sleep quality may help mitigate the effects of poor mental health on social outcomes.

### Moderated Mediation by Environmental Adversity

Environmental moderation analyses revealed that the pathways described above were not uniform across youth. For example, for academic functioning models, financial adversity was a significant moderator in that higher financial adversity weakened associations between sleep quality and the academic outcome in these models. This may suggest that, in the context of financial strain, lifestyle improvements may confer smaller academic functioning benefits. This may be because economic stressors more directly affect academic functioning (e.g., through resource limitations, unstable housing), overshadowing lifestyle-related influences (23), although this hypothesis requires further investigation.

For social problems, the pattern of moderated mediation differed from that observed for models examining academic functioning. Whereas financial adversity weakened lifestyle-related mediation for academic functioning, the opposite was true for social problems: as family conflict and financial adversity increased and school environment worsened, the mediating effects of sleep quality on associations between mental health and social problems strengthened. One possibility is that, under high-conflict or high-adversity conditions, sleep disruptions may exacerbate emotional reactivity, irritability, or social withdrawal, thereby magnifying their downstream impact on social functioning (5, 37). Taken together, these findings indicate that healthy lifestyle mechanisms operate differently across functional domains, with sleep quality becoming a stronger mediator of social functioning under higher adversity, in contrast to the diminished mediation observed for academic functioning.

### Implications for Prevention and Intervention

This study identifies sleep quality and to a lesser extent screen time as potentially tractable intervention targets for improving outcomes among adolescents experiencing mental health concerns. Sleep interventions, such as school-based sleep hygiene programs to improve sleep quality, may meaningfully improve academic and social functioning (38, 39). Yet the moderating effects of financial adversity and family conflict suggest that lifestyle-focused interventions should be tailored to the youth’s environmental context. For youth experiencing economic hardship or high family conflict, sleep-based supports may need to be paired with additional structural supports (e.g., resource assistance, family-based therapies) to maximize impact.

### Limitations

Despite the strengths of the present study, several limitations warrant consideration. First, although the present work focused on modifiable lifestyle variables, it is likely that other lifestyle factors may influence both mental health as well as social and academic functioning. Second, even though mediation effects were statistically robust, several of the effect sizes were modest and may not generalize uniformly across developmental periods. Third, while sleep problems are a diagnostic feature of several mental health conditions (e.g., mood disorders), notably, sleep disturbances partially mediated associations with a range of mental health symptoms, suggesting sleep may function as a separatable and potentially modifiable factor, rather than intrinsic to experiencing specific symptoms such as depression. Finally, although mediation models incorporated independent variables, mediators, and outcomes in chronological order (except for diet, as this information was only available for 1-year follow-up (and would be unlikely to change by 2-year follow-up (25))), this does not necessarily imply that these relationships were causal. Future studies using more sophisticated structural equation models incorporating data from multiple timepoints are necessary to examine the longitudinal relationships between these factors in more comprehensively, and intervention studies are necessary to firmly establish cause-and-effect relationships.

This may necessitate using data from future ABCD studies, as later releases will include more timepoints (including data during middle and late adolescence, when mental issues become increasingly prevalent (40)).

## Conclusions

Findings from this large, multi-site study provide novel evidence that lifestyle factors, especially sleep quality, are key factors linking adolescent mental health symptom domains to academic and social functioning. These pathways are sensitive to contextual conditions, with financial adversity and family conflict moderating the impact lifestyle-related mediation. Together, these results underscore the importance of integrating lifestyle-based and family-informed approaches to support adolescents experiencing early mental health concerns.

## Data Availability

Data used in the preparation of this article were obtained from the Adolescent Brain Cognitive DevelopmentSM (ABCD) Study (https://abcdstudy.org), held in the NIMH Data Archive (NDA).

## Acknowledgements

Dr. Smucny is supported by grant K01-MH125906 and Dr. Karcher by grant R01-MH139880 from the National Institutes of Mental Health.

Data used in the preparation of this article were obtained from the Adolescent Brain Cognitive Development^SM^ (ABCD) Study (https://abcdstudy.org) release 5.1 (DOI: 10.15154/z563-zd24), held in the NIMH Data Archive (NDA). This is a multisite, longitudinal study designed to recruit more than 10,000 children ages 9-10 and follow them over 10 years into early adulthood. The ABCD Study® is supported by the National Institutes of Health and additional federal partners under award numbers U01DA041048, U01DA050989, U01DA051016, U01DA041022, U01DA051018, U01DA051037, U01DA050987, U01DA041174, U01DA041106, U01DA041117, U01DA041028, U01DA041134, U01DA050988, U01DA051039, U01DA041156, U01DA041025, U01DA041120, U01DA051038, U01DA041148, U01DA041093, U01DA041089, U24DA041123, U24DA041147. A full list of supporters is available at https://abcdstudy.org/federal-partners.html. A listing of participating sites and a complete listing of the study investigators can be found at https://abcdstudy.org/consortium_members/. ABCD consortium investigators designed and implemented the study and/or provided data but did not necessarily participate in the analysis or writing of this report. This manuscript reflects the views of the authors and may not reflect the opinions or views of the NIH or ABCD consortium investigators.

## Notes

### Competing Interest Statement

The authors have declared no competing interest.

### Summary of Updates

The article has been revised to update to the most recently submitted version.

## References

1. Barch DM, Albaugh MD, Avenevoli S, Chang L, Clark DB, Glantz MD, Hudziak JJ, Jernigan TL, Tapert SF, Yurgelun-Todd D, Alia-Klein N, Potter AS, Paulus MP, Prouty D, Zucker RA, Sher KJ. Demographic, physical and mental health assessments in the adolescent brain and cognitive development study: Rationale and description. Dev Cogn Neurosci. 2018;32:55–66.

2. Carvajal L, Requejo JH, Irwin CE. The Measurement of Mental Health Problems Among Adolescents and Young Adults Throughout the World. J Adolesc Health. 2021;69:361–362.

3. Achenbach TM, Rescorla LA: Manual for the ASEBA School-Age Forms & Profiles. Burlington, VT, University of Vermont, Research Center for Children, Youth and Families; 2001.

4. Karcher NR, Barch DM, Avenevoli S, Savill M, Huber RS, Simon TJ, Leckliter IN, Sher KJ, Loewy RL. Assessment of the Prodromal Questionnaire-Brief Child Version for Measurement of Self-reported Psychoticlike Experiences in Childhood. JAMA Psychiatry. 2018;75:853–861.

5. Achterbergh L, Pitman A, Birken M, Pearce E, Sno H, Johnson S. The experience of loneliness among young people with depression: a qualitative meta-synthesis of the literature. BMC Psychiatry. 2020;20:415.

6. Dalforno RW, Wengert HI, Kim LP, Jacobsen KH. Anxiety and school absenteeism without permission among adolescents in 69 low-and middle-income countries. Dialogues Health. 2022;1:100046.

7. Finning K, Ukoumunne OC, Ford T, Danielsson-Waters E, Shaw L, Romero De Jager I, Stentiford L, Moore DA. The association between child and adolescent depression and poor attendance at school: A systematic review and meta-analysis. J Affect Disord. 2019;245:928–938.

8. Kearney CA, Dupont R, Fensken M, Gonzálvez C. School attendance problems and absenteeism as early warning signals: review and implications for health-based protocols and school-based practices. Frontiers in Education. 2023;Volume 8 - 2023.

9. Kupferberg A, Hasler G. The social cost of depression: Investigating the impact of impaired social emotion regulation, social cognition, and interpersonal behavior on social functioning. Journal of Affective Disorders Reports. 2023;14:100631.

10. Huckle C, Lemmel F, Johnson S. Experiences of friendships of young people with first-episode psychosis: A qualitative study. PLoS One. 2021;16:e0255469.

11. Steenkamp LR, Bolhuis K, Blanken LME, Luijk M, Hillegers MHJ, Kushner SA, Tiemeier H. Psychotic experiences and future school performance in childhood: a population-based cohort study. J Child Psychol Psychiatry. 2021;62:357–365.

12. Firth J, Solmi M, Wootton RE, Vancampfort D, Schuch FB, Hoare E, Gilbody S, Torous J, Teasdale SB, Jackson SE, Smith L, Eaton M, Jacka FN, Veronese N, Marx W, Ashdown-Franks G, Siskind D, Sarris J, Rosenbaum S, Carvalho AF, Stubbs B. A meta-review of “lifestyle psychiatry”: the role of exercise, smoking, diet and sleep in the prevention and treatment of mental disorders. World Psychiatry. 2020;19:360–380.

13. Ruiz-Ranz E, Asin-Izquierdo I. Physical activity, exercise, and mental health of healthy adolescents: A review of the last 5 years. Sports Med Health Sci. 2025;7:161–172.

14. Bruni O, Ottaviano S, Guidetti V, Romoli M, Innocenzi M, Cortesi F, Giannotti F. The Sleep Disturbance Scale for Children (SDSC). Construction and validation of an instrument to evaluate sleep disturbances in childhood and adolescence. J Sleep Res. 1996;5:251–261.

15. Nagata JM, Bashir A, Weinstein S, Al-Shoaibi AAA, Shao IY, Ganson KT, Testa A, Garber AK. Social epidemiology of the Mediterranean-dietary approaches to stop hypertension intervention for neurodegenerative delay (MIND) diet among early adolescents: the Adolescent Brain Cognitive Development Study. Pediatr Res. 2024;96:230–236.

16. Camprodon-Boadas P, Gil-Dominguez A, De la Serna E, Sugranyes G, Lazaro I, Baeza I. Mediterranean Diet and Mental Health in Children and Adolescents: A Systematic Review. Nutr Rev. 2025;83:e343–e355.

17. Santos RMS, Mendes CG, Sen Bressani GY, de Alcantara Ventura S, de Almeida Nogueira YJ, de Miranda DM, Romano-Silva MA. The associations between screen time and mental health in adolescents: a systematic review. BMC Psychology. 2023;11:127.

18. Zablotsky B, Ng AE, Black LI, Haile G, Bose J, Jones JR, Blumberg SJ. Associations Between Screen Time Use and Health Outcomes Among US Teenagers. Prev Chronic Dis. 2025;22:E38.

19. Zink J, O’Connor SG, Blachman-Demner DR, Wolff-Hughes DL, Berrigan D. Examining the Bidirectional Associations Between Sleep Duration, Screen Time, and Internalizing Symptoms in the ABCD Study. J Adolesc Health. 2024;74:496–503.

20. Reeve S, Bell V. Sleep disorders predict the 1-year onset, persistence, but not remission of psychotic experiences in preadolescence: a longitudinal analysis of the ABCD cohort data. Eur Child Adolesc Psychiatry. 2023;32:1609–1619.

21. Damme KSF, Vargas TG, Walther S, Shankman SA, Mittal VA. Physical and mental health in adolescence: novel insights from a transdiagnostic examination of FitBit data in the ABCD study. Transl Psychiatry. 2024;14:75.

22. Aliyas Z, Mahmoudian A, Cloutier M-S. Association Between Socio-Emotional Health, Physical Activity and Screen Time Among Children. American Journal of Health Education. 2025;56:366–375.

23. Gautam N, Rahman MM, Khanam R. Socioeconomic inequalities in health behaviors in children and adolescents: evidence from an Australian cohort. BMC Public Health. 2025;25:314.

24. Al-Shoaibi AAA, Zamora G, Chu J, Patel KP, Ganson KT, Testa A, Jackson DB, Tapert SF, Baker FC, Nagata JM. Family conflict and less parental monitoring were associated with greater screen time in early adolescence. Acta Paediatr. 2024;113:2452–2458.

25. da Costa MP, Severo M, Araujo J, Vilela S. Longitudinal tracking of diet quality from childhood to adolescence: The Interplay of individual and sociodemographic factors. Appetite. 2024;196:107279.

26. Achenbach TM, Ruffle TM. The Child Behavior Checklist and related forms for assessing behavioral/emotional problems and competencies. Pediatr Rev. 2000;21:265–271.

27. Echeverria SE, Diez-Roux AV, Link BG. Reliability of self-reported neighborhood characteristics. J Urban Health. 2004;81:682–701.

28. Moos RH: Family Environment Scale Manual: Development, Applications, Research, Consult. Psychol. Press; 1994.

29. Zucker RA, Gonzalez R, Feldstein Ewing SW, Paulus MP, Arroyo J, Fuligni A, Morris AS, Sanchez M, Wills T. Assessment of culture and environment in the Adolescent Brain and Cognitive Development Study: Rationale, description of measures, and early data. Dev Cogn Neurosci. 2018;32:107–120.

30. Karcher NR, Loewy RL, Savill M, Avenevoli S, Huber RS, Simon TJ, Leckliter IN, Sher KJ, Barch DM. Replication of Associations With Psychotic-Like Experiences in Middle Childhood From the Adolescent Brain Cognitive Development (ABCD) Study. Schizophr Bull Open. 2020;1:sgaa009.

31. Luciana M, Bjork JM, Nagel BJ, Barch DM, Gonzalez R, Nixon SJ, Banich MT. Adolescent neurocognitive development and impacts of substance use: Overview of the adolescent brain cognitive development (ABCD) baseline neurocognition battery. Dev Cogn Neurosci. 2018;32:67–79.

32. Roth B, Becker N, Romeyke S, Schäfer S, Domnick F, Spinath FM. Intelligence and school grades: A meta-analysis. Intelligence. 2015;53:118–137.

33. Almat NS, Aliya MS, Zhanna UT, Gulmira DS. The Relationship Between Social Intelligence And IQ: A Psychometric Analysis. The Open Psychology Journal. 2023;16.

34. Finning K, Ukoumunne OC, Ford T, Danielson-Waters E, Shaw L, Romero De Jager I, Stentiford L, Moore DA. Review: The association between anxiety and poor attendance at school - a systematic review. Child Adolesc Ment Health. 2019;24:205–216.

35. Qiu J, Morales-Munoz I. Associations between Sleep and Mental Health in Adolescents: Results from the UK Millennium Cohort Study. Int J Environ Res Public Health. 2022;19.

36. Woodfield M, Butler NG, Tsappis M. Impact of sleep and mental health in adolescence: an overview. Curr Opin Pediatr. 2024;36:375–381.

37. Cummings EM, Koss KJ, Davies PT. Prospective relations between family conflict and adolescent maladjustment: security in the family system as a mediating process. J Abnorm Child Psychol. 2015;43:503–515.

38. Fenwick-Smith A, Dahlberg EE, Thompson SC. Systematic review of resilience-enhancing, universal, primary school-based mental health promotion programs. BMC Psychol. 2018;6:30.

39. Green JG, McLaughlin KA, Alegria M, Costello EJ, Gruber MJ, Hoagwood K, Leaf PJ, Olin S, Sampson NA, Kessler RC. School mental health resources and adolescent mental health service use. J Am Acad Child Adolesc Psychiatry. 2013;52:501–510.

40. Substance Abuse and Mental Health Services Administration (2013). Behavioral Health, United States, 2012. HHS Publication No. (SMA) 13-4797. Rockville, MD: Substance Abuse and Mental Health Services Administration.

