## Supplementary Information for "Mediating Effects of Healthy Lifestyle Factors on Associations between Mental Health and Functional Outcomes in Early Adolescence"

**Supplementary Methods**

***Participant Exclusion Criteria***

Potential participants were excluded from ABCD study participation for the following reasons: child not fluent in English, major neurological disorder, gestational age less than 28 weeks or birthweight less than 1,200 grams, history of traumatic brain injury, or has a current diagnosis of schizophrenia, autism spectrum disorder (moderate, severe), mental retardation/intellectual disability, or alcohol/substance use disorder. For families with siblings, only one sibling from the family was included (based on alphabetical order from the Global Unique Identifier).

***Instruments***

*Mediterranean-DASH Intervention for Neurodegenerative Delay (MIND)*

The MIND diet questionnaire is a 15-item questionnaire that assesses intake frequencies for whole grains, vegetables, leafy vegetables, berries, nuts, poultry, beans, fish, wine, and olive oil. It also asks about the intake of foods that have been associated with higher risk of neurodegeneration, such as fast and fried foods, pastries and sweets, butter, and cheese. The assessment given by the ABCD study omitted the category of wine, leaving 14 categories of food. The survey asks whether the child consumed a specific serving size of each food group weekly over the last year ("yes" = 1, "no" = 0). The total score of the 14 MIND questions was then calculated and used as a healthy lifestyle mediator in analyses. As described in (1), because children are more likely to inaccurately report food records (2-4) MIND data was taken from parental report.

*Sleep Disturbance Scale for Children (SDSC)*

The SDSC is a 26 item, Likert-style questionnaire designed to evaluate sleep disorders in children and provide an overall measure of sleep disturbance suitable for use in clinical screening and research (5). Most questions on the scale are rated in terms of frequency from 1 (never) to 5 (always) and summed to yield a composite score (one question is rated 1-5 based on how many hours the child sleeps each night, and the another is rated 1-5 based on how long it takes for the child to fall asleep). Example questions include "the child has difficulty getting to sleep each night" and "the child wakes up more than twice per night." Scores were reverse coded such that higher scores corresponded to better sleep quality.

*ABCD Neighborhood Crime and Safety Survey*

The ABCD Neighborhood Crime and Safety Survey (6) includes three items ("My neighborhood is safe from crime; Violence is not a problem in my neighborhood; I feel safe walking in my neighborhood, day or night") ranked from 1 to 5 ("strongly agree" to "strongly disagree"). These items were summed to yield a composite score, with lower scores corresponding to safer neighborhoods.

*ABCD Parent Family Environment Scale-Family Conflict Subscale (Modified from PhenX)*

The Family Conflict subscale (7) is a nine-item instrument made up of yes/no questions (coded 0 or 1) that are summed together to yield a composite score. Questions include "family members rarely become opening angry" and "family members sometimes hit each other." Higher scores indicate more family conflict.

*School Risk and Protective Factors School Environment Subscale (Modified from PhenX)*

The School Environment subscale (8) is a 12-item questionnaire with 4 possible answers per question (1 = NO!; 2 = no; 3 = yes; 4 = YES!). Example questions include "In my school, students have lots of chances to help decide things like class activities and rules" and "I get along with my teachers." Higher scores indicate better school environment.

*Family Financial Adversity*

As described in (9), financial adversity was measured as the summed endorsement of 7 parent-reported questions of financial difficulties experienced during the past 12 months from a demographic questionnaire. This measure included the following questions (participants received a score of 1 for each endorsed item, and financial adversity was scored as the sum of these endorsed items): (1) Needed food but couldn’t afford to buy it or couldn’t afford to go out to get it?, (2) Were without telephone service because you could not afford it?, (3) Didn’t pay the full amount of the rent or mortgage because you could not afford it?, (4) Were evicted from your home for not paying the rent or mortgage?, (5) Had services turned off by the gas or electric company, or the oil company wouldn’t deliver oil because payments were not made?, (6) Had someone who needed to see a doctor or go to the hospital but didn’t go because you could not afford it?, and (7) Had someone who needed a dentist but couldn’t go because you could not afford it?

***Mediation Analyses: Statistics***

Significance of mediation models was determined by calculating the 95% confidence interval (CI) of the indirect effect based on 5000 bias-corrected bootstrapped samples; an indirect effect in which the corresponding 95% CI was entirely positive or negative implies that *p* < .05. Percent mediation was also calculated using the indirect effect for each individual mediator as well as across all mediators combined by the equation *indirect effect* ÷ *total effect* × 100. Significant indirect effects were only considered to be of interest if the total effect (between the independent and dependent variable, including the mediator) was significant (*p* < .05).

***Moderated Mediation Effects***

Effects of environmental moderators were examined using PROCESS v.5.0. A diagram of the analysis strategy for environmental moderation effects is shown in Supplementary Figure 1. These moderated mediation models examined the degree to which each moderator (e.g., family financial adversity) affected the mediation of associations between mental health with outcome by affecting the relationships between lifestyle factors and outcome (i.e., the mediation model "b" path). We examined moderation of the "b" path (i.e., the path linking lifestyle factors to outcome), as the goal of this study was to examine everyday lifestyle mediators (e.g., screen time habits) that can be altered independent of a clinical provider. Concordantly, we were particularly interested to examine how the beneficial effects of these mediators might be influenced by living conditions. Similar to the primary (non-moderated) mediation analyses, significant moderated mediation was determined by calculating the 95% confidence interval (CI) of the index of moderated mediation based on 5000 bias-corrected bootstrapped samples.

**Supplementary Results**

*Mediation Analyses: Mental Health Predicting School Grades*

Including cognition as a covariate did not appreciably alter results of mediation models with academic functioning as the outcome (Supplementary Figure 2, Supplementary Table 2). When weekday screen time was used instead of weekend screen time, the mediating effects of screen time were consistently smaller (e.g., 3.7% (weekday) vs. 6.3% (weekend) for depression), although it remained statistically significant for all four psychopathological domains tested (Supplementary Figure 3, Supplementary Table 3).

*Mediation Analyses: Mental Health Predicting Social Functioning*

Including cognition as a covariate did not appreciably alter results of mediation models with social functioning as the outcome (Supplementary Figure 4, Supplementary Table 5). When weekday screen time was used instead of weekend screen time, the mediating effects of screen time became consistently smaller and non-significant for all psychopathological domains tested (Supplementary Figure 5, Supplementary Table 6).

**Supplementary Table 1.** Results from mediation analyses in which Child Behavioral Checklist (CBCL) depression, anxiety, or total problems t-scores or Prodromal Questionnaire-Brief Child (PQ-BC) version distress score at 1-year follow-up were used as predictors (independent variables), healthy lifestyle indicators (sleep quality (reverse score of the Sleep Disturbance Scale for Children, year 2), weekend screen time (reverse scored, year 2), physical activity (year 2), and Mediterranean diet (year 1)) as mediators, and academic functioning (grades between follow-up years 2-3) as the outcome (dependent variable). Covariates included age (year 2), sex, and site. %Mediation is calculated as Indirect Effect ÷ Total Effect × 100. **p* < .05. Mediators are listed in order of decreasing %mediation. Abbreviations: CI = Confidence Interval, SE = Standard Error.

| **Predictor** | **Predictor to Mediator B (SE, t, *p*)** | **Mediator to Outcome B (SE, t, *p*)** | **Total Effect, i.e., Predictor + Mediators to Outcome B (SE, t, *p*)** | **Direct Effect, i.e., Predictor to Outcome B (SE, t, *p*)** | **Indirect Effect, i.e., Mediation B (SE, Bootstrapped 95% CI)** | **%Mediation If Total and Indirect Effects Significant** |
| --- | --- | --- | --- | --- | --- | --- |
| **Mediator** |  |  |  |  |  |  |
| **CBCL Depression** | – | – | -.0682 (.0046, -14.84, <.001)* | -.0481 (.0051, -9.42, <.001)* | – | – |
| **Sleep Quality** | -.5945 (.0146, -40.63, <.001)* | .0212 (.0038, 5.61, <.001)* | – | – | -.0126 (.0026, -.0177 – -.0076)* | 18.5% |
| **Weekend Screen Time (Reverse-Scored)** | -.0520 (.0062, -8.39, <.001)* | .0831 (.0091, 9.11, <.001)* | – | – | -.0043 (.0008, -.0060 – -.0030)* | 6.3% |
| **Physical Activity** | -.0246 (.0044, -5.65, <.001)* | .0770 (.0128, 6.02, <.001)* | – | – | -.0019 (.0005, -.0030 – -.0011)* | 2.8% |
| **Diet** | -.0650 (.0048, -13.52, <.001)* | .0190 (.0116, 1.63, .10) | – | – | -.0012 (.0008, -.0028 – .0004)* | 1.8% (n.s.) |
| **All Mediators** | – | – | – | – | -.0201 (.0028, -.0256 – -.0148)* | 30% |
| **CBCL Anxiety** | – | – | -.0402 (.0045, -8.83, <.001)* | -.0209 (.0048, -4.35, <.001)* | – | – |
| **Sleep Quality** | -.4716 (.0149, -31.61, <.001)* | .0309 (.0037, 8.43, <.001)* | – | – | -.0146 (.0021, -.0189 – -.0107)* | 36.3% |
| **Weekend Screen Time (Reverse-Scored)** | -.0238 (.0061, -3.90, <.001)* | .0859 (.0092, 9.37, <.001)* | – | – | -.0020 (.0006, -.0034 – -.0010)* | 5.0% |
| **Physical Activity** | -.0202 (.0043, -4.72, <.001)* | .0802 (.0129, 6.24, <.001)* | – | – | -.0016 (.0005, -.0027 – -.0008)* | 4.0% |
| **Diet** | -.0356 (.0048, -7.48, <.001)* | .0296 (.0116, 2.55, .011)* | – | – | -.0011 (.0005, -.0020 – .0002)* | 2.7% |
| **All Mediators** | – | – | – | – | -.0193 (.0023, -.0237 – -.0148)* | 48% |
| **PQ-BC Distress** | – | – | -.0372 (.0031, -12.21, <.001)* | -.0305 (.0030, -10.09, <.001)* | – | – |
| **Sleep Quality** | -.0942 (.0107, -8.79, <.001)* | .0330 (.0034, 9.63, <.001)* | – | – | -.0031 (.0006, -.0044 – -.0021)* | 8.3% |
| **Weekend Screen Time (Reverse-Scored)** | -.0281 (.0041, -6.84, <.001)* | .0809 (.0091, 8.89, <.001)* | – | – | -.0023 (.0005, -.0033 – -.0014)* | 6.2% |
| **Physical Activity** | -.0172 (.0029, -5.99, <.001)* | .0740 (.0128, 5.79, <.001)* | – | – | -.0013 (.0003, -.0020 – -.0007)* | 3.5% |
| **Diet** | -.0022 (.0032, -1.51, .13)* | .0337 (.0115, 2.92, .004)* | – | – | -.0001 (.0001, -.0004 – .0002) | .3% (n.s.) |
| **All Mediators** | – | – | – | – | -.0067 (.0008, -.0085 – -.0052)* | 18% |
| **CBCL Total Problems** | – | – | -.0565 (.0025, -22.98, <.001)* | -.0508 (.0028, -18.26, <.001)* | – | – |
| **Weekend Screen Time (Reverse-Scored)** | -.0328 (.0034, -9.70, <.001)* | .0785 (.0090, 8.75, <.001)* | – | – | -.0026 (.0004, -.0035 – -.0018)* | 4.6% |
| **Sleep Quality** | -.3400 (.0079, -43.05, <.001)* | .0056 (.0038, 1.49, .14) | – | – | -.0019 (.0014, -.0047 – .0009) | 3.4% (n.s.) |
| **Physical Activity** | -.0137 (.0024, -5.76, <.001)* | .0721 (.0126, 5.74, <.001)* | – | – | -.0010 (.0003, -.0016 – -.0006)* | 1.8% |
| **Diet** | -.0371 (.0026, -14.15, <.001)* | .0056 (.0114, .49, .62) | – | – | -.0002 (.0005, -.0011 – .0007) | .4% (n.s.) |
| **All Mediators** | – | – | – | – | -.0057 (.0015, -.0087 – -.0027)* | 10% |

**Supplementary Table 2.** Results from mediation analyses in which Child Behavioral Checklist (CBCL) depression, anxiety, or total problems t-scores or Prodromal Questionnaire-Brief Child (PQ-BC) version distress score at 1-year follow-up were used as predictors (independent variables), healthy lifestyle indicators (sleep quality (reverse score of the Sleep Disturbance Scale for Children, year 2), weekend screen time (reverse scored, year 2), physical activity (year 2), and Mediterranean diet (year 1)) as mediators, and academic functioning (grades between follow-up years 2-3) as the outcome (dependent variable). Covariates included age (year 2), sex, site, and NIH Cognition Toolbox Composite raw score. %Mediation is calculated as Indirect Effect ÷ Total Effect × 100. **p* < .05. Mediators are listed in order of decreasing %mediation. Abbreviations: CI = Confidence Interval, n.s. = non-significant, SE = Standard Error.

| **Predictor** | **Predictor to Mediator B (SE, t, *p*)** | **Mediator to Outcome B (SE, t, *p*)** | **Total Effect, i.e., Predictor + Mediators to Outcome B (SE, t, *p*)** | **Direct Effect, i.e., Predictor to Outcome B (SE, t, *p*)** | **Indirect Effect, i.e., Mediation B (SE, Bootstrapped 95% CI)** | **%Mediation** |
| --- | --- | --- | --- | --- | --- | --- |
| **Mediator** |  |  |  |  |  |  |
| **CBCL Depression** | – | – | -.0612 (.0043, -14.36, <.001)* | -.0413 (.0048, -8.68, <.001)* | – | – |
| **Sleep Quality** | -.5938 (.0147, -40.34, <.001)* | .0223 (.0035, 6.29, <.001)* | – | – | -.0132 (.0024, -.0181 – -.0085)* | 21.6% |
| **Weekend Screen Time (Reverse-Scored)** | -.0495 (.0062, -7.98, <.001)* | .0575 (.0086, 6.71, <.001)* | – | – | -.0029 (.0006, -.0043 – -.0018)* | 4.7% |
| **Physical Activity** | -.0237 (.0044, -5.43, <.001)* | .0431 (.0120, 3.60, .003)* | – | – | -.0010 (.0004, -.0019 – -.0005)* | 4.6% |
| **Diet** | -.0665 (.0048, -13.75, <.001)* | .0426 (.0109, 3.91, <.001)* | – | – | -.0028 (.0008, -.0046 – -.0013)* | 1.6% |
| **All Mediators** | – | – | – | – | -.0199 (.0025, -.0251 – -.0150)* | 33% |
| **CBCL Anxiety** | – | – | -.0355 (.0042, -8.41, <.001)* | -.0168 (.0045, -3.75, <.001)* | – | – |
| **Sleep Quality** | -.4739 (.0150, -31.52, <.001)* | .0310 (.0034, 9.03, <.001)* | – | – | -.0147 (.0020, -.0187 – -.0109)* | 41.4% |
| **Weekend Screen Time (Reverse-Scored)** | -.0228 (.0061, -3.74, <.001)* | .0596 (.0086, 6.92, <.001)* | – | – | -.0014 (.0005, -.0025 – -.0006)* | 5.4% |
| **Physical Activity** | -.0185 (.0043, -4.32, <.001)* | .0461 (.0120, 3.83, <.001)* | – | – | -.0009 (.0003, -.0016 – -.0003)* | 3.9% |
| **Diet** | -.0359 (.0048, -7.49, <.001)* | .0523 (.0109, 4.81, <.001)* | – | – | -.0019 (.0005, -.0030 – .0010)* | 2.5% |
| **All Mediators** | – | – | – | – | -.0188 (.0021, -.0230 – -.0147)* | 53% |
| **PQ-BC Distress** | – | – | -.0224 (.0029, -7.70, <.001)* | -.0169 (.0029, -5.89, <.001)* | – | – |
| **Sleep Quality** | -.0953 (.0110, -8.66, <.001)* | .0335 (.0032, 10.42, <.001)* | – | – | -.0032 (.0006, -.0045 – -.0022)* | 14.3% |
| **Weekend Screen Time (Reverse-Scored)** | -.0234 (.0042, -5.57, <.001)* | .0576 (.0086, 6.71, <.001)* | – | – | -.0014 (.0004, -.0022 – -.0008)* | 6.3% |
| **Physical Activity** | -.0142 (.0029, -4.83, <.001)* | .0437 (.0120, 3.63, .<001)* | – | – | -.0006 (.0002, -.0012 – -.0003)* | 2.7% |
| **Diet** | -.0047 (.0033, -1.42, .16) | .0544 (.0108, 5.02, <.001)* | – | – | -.0003 (.0002 -.0007 – .0001) | 1.3% (n.s.) |
| **All Mediators** | – | – | – | – | -.0054 (.0008, -.0071 – -.0040)* | 24% |
| **CBCL Total Problems** | – | – | -.0482 (.0023, -20.93, <.001)* | -.0414 (.0026, -15.81, <.001)* | – | – |
| **Sleep Quality** | -.3424 (.0080, -42.87, <.001)* | .0102 (.0035, 2.89, .004)* | – | – | -.0035 (.0014, -.0063 – -.0009)* | 7.3% |
| **Weekend Screen Time (Reverse-Scored)** | -.0299 (.0034, -8.80, <.001)* | .0551 (.0085, 6.51, <.001)* | – | – | -.0017 (.0004, -.0024 – -.0010)* | 3.5% |
| **Diet** | -.0386 (.0027, -14.56, <.001)* | .0314 (.0108, 2.92, .003)* | – | – | -.0012 (.0005, -.0021 – .0004)* | 2.4% |
| **Physical Activity** | -.0118 (.0024, -4.93, <.001)* | .0409 (.0118, 3.46, <.001)* | – | – | -.0005 (.0002, -.0009 – -.0002)* | 1.0% |
| **All Mediators** | – | – | – | – | -.0068 (.0014, -.0098 – -.0042)* | 14.1% |

**Supplementary Table 3.** Results from mediation analyses in which Child Behavioral Checklist (CBCL) depression, anxiety, or total problems t-scores or Prodromal Questionnaire-Brief Child (PQ-BC) version distress score at 1-year follow-up were used as predictors (independent variables), healthy lifestyle indicators (sleep quality (reverse score of the Sleep Disturbance Scale for Children, year 2), weekday screen time (reverse scored, year 2), physical activity (year 2), and Mediterranean diet (year 1)) as mediators, and academic functioning (grades between follow-up years 2-3) as the outcome (dependent variable). Covariates included age (year 2), sex, and site. %Mediation is calculated as Indirect Effect ÷ Total Effect × 100. **p* < .05. Mediators are listed in order of decreasing %mediation. Abbreviations: CI = Confidence Interval, n.s. = non-significant, SE = Standard Error.

| **Predictor** | **Predictor to Mediator B (SE, t, *p*)** | **Mediator to Outcome B (SE, t, *p*)** | **Total Effect, i.e., Predictor + Mediators to Outcome B (SE, t, *p*)** | **Direct Effect, i.e., Predictor to Outcome B (SE, t, *p*)** | **Indirect Effect, i.e., Mediation B (SE, Bootstrapped 95% CI)** | **%Mediation** |
| --- | --- | --- | --- | --- | --- | --- |
| **Mediator** |  |  |  |  |  |  |
| **CBCL Depression** | – | – | -.0682 (.0046, -14.84, <.001)* | -.0486 (.0051, -9.52, <.001)* | – | – |
| **Sleep Quality** | -.5945 (.0146, -40.63, <.001)* | .0224 (.0038, 5.94, <.001)* | – | – | -.0133 (.0026, -.0187 – -.0084)* | 19.5% |
| **Weekday Screen Time (Reverse-Scored)** | -.0297 (.0063, -4.74, <.001)* | .0858 (.0089, 9.64, <.001)* | – | – | -.0025 (.0007, -.0040 – -.0014)* | 3.7% |
| **Physical Activity** | -.0246 (.0044, -5.65, <.001)* | .0797 (.0127, 6.25, <.001)* | – | – | -.0020 (.0005, -.0031 – -.0011)* | 2.9% |
| **Diet** | -.0650 (.0048, -13.52, <.001)* | .0271 (.0115, 2.36, .019)* | – | – | -.0018 (.0008, -.0034 – -.0002)* | 2.6% |
| **All Mediators** | – | – | – | – | -.0196 (.0028, -.0255 – -.0143)* | 29% |
| **CBCL Anxiety** | – | – | -.0402 (.0045, -8.83, <.001)* | -.0206 (.0048, -4.30, <.001)* | – | – |
| **Sleep Quality** | -.4716 (.0149, -31.61, <.001)* | .0324 (.0037, 8.87, <.001)* | – | – | -.0153 (.0021, -.0196 – -.0114)* | 38.0% |
| **Physical Activity** | -.0202 (.0043, -4.72, <.001)* | .0832 (.0128, 6.50, <.001)* | – | – | -.0017 (.0005, -.0027 – -.0009)* | 4.2% |
| **Diet** | -.0356 (.0048, -7.48, <.001)* | .0383 (.0115, 3.33, <.001)* | – | – | -.0014 (.0005, -.0024 – .0005)* | 3.5% |
| **Weekday Screen Time (Reverse-Scored)** | -.0139 (.0061, -2.27, .023)* | .0874 (.0089, 9.76, <.001)* | – | – | -.0012 (.0006, -.0024 – -.0001)* | 3.0% |
| **All Mediators** | – | – | – | – | -.0195 (.0023, -.0242 – -.0154)* | 48.5% |
| **PQ-BC Distress** | – | – | -.0372 (.0031, -12.21, <.001)* | -.0310 (.0030, -10.27, <.001)* | – | – |
| **Sleep Quality** | -.0942 (.0107, -8.79, <.001)* | .0343 (.0034, 10.03, <.001)* | – | – | -.0032 (.0006, -.0045 – -.0022)* | 8.6% |
| **Weekday Screen Time (Reverse-Scored)** | -.0195 (.0041, -4.73, <.001)* | .0839 (.0089, 9.43, <.001)* | – | – | -.0016 (.0004, -.0025 – -.0010)* | 4.3% |
| **Physical Activity** | -.0172 (.0029, -5.99, <.001)* | .0764 (.0127, 6.00, <.001)* | – | – | -.0013 (.0003, -.0021 – -.0007)* | 3.5% |
| **Diet** | -.0022 (.0032, -.67, .50) | .0417 (.0114, 3.66, <.001)* | – | – | -.0001 (.0002 -.0004 – .0002) | .3% (n.s.) |
| **All Mediators** | – | – | – | – | -.0063 (.0008, -.0081 – -.0049)* | 17% |
| **CBCL Total Problems** | – | – | -.0565 (.0025, -22.98, <.001)* | -.0513 (.0028, -18.50, <.001)* | – | – |
| **Sleep Quality** | -.3400 (.0079, -43.05, <.001)* | .0065 (.0038, 1.72, .084) | – | – | -.0022 (.0015, -.0050 – -.0006) | 3.9% (n.s.) |
| **Weekday Screen Time (Reverse-Scored)** | -.0171 (.0034, -4.99, <.001)* | .0842 (.0087, 9.63, <.001)* | – | – | -.0014 (.0003, -.0022 – -.0008)* | 2.5% |
| **Physical Activity** | -.0137 (.0024, -5.76, <.001)* | .0741 (.0125, 5.92, <.001)* | – | – | -.0010 (.0003, -.0016 – -.0006)* | 1.8% |
| **Diet** | -.0371 (.0026, -14.15, <.001)* | .0129 (.0113, 1.14, .26) | – | – | -.0005 (.0005, -.0014 – .0004) | .9% (n.s.) |
| **All Mediators** | – | – | – | – | -.0051 (.0016, -.0082 – -.0021)* | 9% |

**Supplementary Table 4.** Results from mediation analyses in which Child Behavioral Checklist (CBCL) depression or anxiety t-scores or Prodromal Questionnaire-Brief Child (PQ-BC) version distress score at 1-year follow-up were used as predictors (independent variables), healthy lifestyle indicators (sleep quality (reverse score of the Sleep Disturbance Scale for Children, year 2), weekend screen time (reverse scored, year 2), physical activity (year 2), and Mediterranean diet (year 1)) as mediators, and social problems (CBCL Social Problems score) as the outcome (dependent variable). Covariates included age (year 2), sex, and site. %Mediation is calculated as Indirect Effect ÷ Total Effect × 100. **p* < .05. CBCL Total Problems was not used as a predictor as it includes CBCL social score when calculating its value. Mediators are listed in order of decreasing %mediation. Abbreviations: CI = Confidence Interval, n.s. = non-significant, SE = Standard Error.

| **Predictor** | **Predictor to Mediator B (SE, t, *p*)** | **Mediator to Outcome B (SE, t, *p*)** | **Total Effect, i.e., Predictor + Mediators to Outcome B (SE, t, *p*)** | **Direct Effect, i.e., Predictor to Outcome B (SE, t, *p*)** | **Indirect Effect, i.e., Mediation B (SE, Bootstrapped 95% CI)** | **%Mediation** |
| --- | --- | --- | --- | --- | --- | --- |
| **Mediator** |  |  |  |  |  |  |
| **CBCL Depression** | – | – | .3470 (.0084, 41.44, <.001)* | .2732 (.0092, 29.55, <.001)* | – | – |
| **Sleep Quality** | -.5845 (.0137, -42.67, <.001)* | -.1180 (.0069, -17.05, <.001)* | – | – | .0690 (.0063, .0574 – .0819)* | 19.9% |
| **Physical Activity** | -.0260 (.0041, -6.42, <.001)* | -.0762 (.0235, -3.24, .001)* | – | – | .0020 (.0007, .0008 – .0036)* | .6% |
| **Weekend Screen Time (Reverse-Scored)** | -.0528 (.0059, -8.99, <.001)* | -.0253 (.0165, -1.53, .13) | – | – | .0013 (.0010, -.0006 – .0034) | .4% (n.s.) |
| **Diet** | -.0659 (.0045, -14.59, <.001)* | -.0225 (.0212, -1.06, .29) | – | – | .0015 (.0015, -.0013 – .0045) | .4% (n.s.) |
| **All Mediators** | – | – | – | – | .0738 (.0064, .0620 – .0872)* | 21% |
| **CBCL Anxiety** | – | – | .3213 (.0082, 38.98, <.001)* | .2532 (.0086, 29.51, <.001)* | – | – |
| **Sleep Quality** | -.4595 (.0139, -33.09, <.001)* | -.1370 (.0067, -20.54, <.001)* | – | – | .0630 (.0051, .0531 – .0736)* | 19.6% |
| **Diet** | -.0379 (.0044, -8.53, <.001)* | -.0617 (.0211, -2.92, .004)* | – | – | .0023 (.0009, .0007 – .0041)* | .7% |
| **Physical Activity** | -.0210 (.0040, -5.31, <.001)* | -.0798 (.0235, -3.40, <.001)* | – | – | .0017 (.0006, .0006 – .0031)* | .5% |
| **Weekend Screen Time (Reverse-Scored)** | -.0263 (.0057, -4.58, .023)* | -.0420 (.0165, -2.55, .011)* | – | – | .0011 (.0006, .0002 – .0026)* | .3% |
| **All Mediators** | – | – | – | – | .0681 (.0052, .0582 – .0787)* | 21% |
| **PQ-BC Distress** | – | – | .0808 (.0061, 13.15, <.001)* | .0592 (.0058, 10.16, <.001)* | – | – |
| **Sleep Quality** | -.0947 (.0102, -9.32, <.001)* | -.1989 (.0066, -30.15, <.001)* | – | – | .0188 (.0027, .0140 – .0247)* | 23.3% |
| **Physical Activity** | -.0174 (.0027, -6.39, <.001)* | -.0981 (.0247, -3.98, <.001)* | – | – | .0017 (.0005, .0008 – .0029)* | 2.1% |
| **Weekend Screen Time (Reverse-Scored)** | -.0281 (.0039, -7.12, <.001)* | -.0282 (.0173, -1.63, .10) | – | – | .0008 (.0006, -.0002 – .0020)* | 1.0% |
| **Diet** | -.0029 (.0031, -.93, .35) | -.1042 (.0221, -4.72, <.001)* | – | – | .0003 (.0004, -.0004 – .0011)* | .4% |
| **All Mediators** | – | – | – | – | .0216 (.0028, .0166 – .0277)* | 27% |

**Supplementary Table 5.** Results from mediation analyses in which Child Behavioral Checklist (CBCL) depression or anxiety t-scores or Prodromal Questionnaire-Brief Child (PQ-BC) version distress score at 1-year follow-up were used as predictors (independent variables), healthy lifestyle indicators (sleep quality (reverse score of the Sleep Disturbance Scale for Children, year 2), weekend screen time (reverse scored, year 2), physical activity (year 2), and Mediterranean diet (year 1)) as mediators, and social problems (CBCL Social Problems score) as the outcome (dependent variable). Covariates included age (year 2), sex, site, and NIH Cognition Toolbox Composite raw score. %Mediation is calculated as Indirect Effect ÷ Total Effect × 100. **p* < .05. CBCL Total Problems was not used as a predictor as it includes CBCL social score when calculating its value. Mediators are listed in order of decreasing %mediation. Abbreviations: CI = Confidence Interval, n.s. = non-significant, SE = Standard Error.

| **Predictor** | **Predictor to Mediator B (SE, t, *p*)** | **Mediator to Outcome B (SE, t, *p*)** | **Total Effect, i.e., Predictor + Mediators to Outcome B (SE, t, *p*)** | **Direct Effect, i.e., Predictor to Outcome B (SE, t, *p*)** | **Indirect Effect, i.e., Mediation B (SE, Bootstrapped 95% CI)** | **%Mediation** |
| --- | --- | --- | --- | --- | --- | --- |
| **Mediator** |  |  |  |  |  |  |
| **CBCL Depression** | – | – | .3451 (.0083, 41.41, <.001)* | .2739 (.0092, 29.70, <.001)* | – | – |
| **Sleep Quality** | -.5849 (.0138, -42.44, <.001)* | -.1153 (.0069, -16.65, <.001)* | – | – | .0674 (.0061, .0560 – .0797)* | 19.5% |
| **Diet** | -.0671 (.0046, -14.75, <.001)* | -.0297 (.0212, -1.40, .16) | – | – | .0020 (.0015, -.0008 – .0050) | .6% (n.s.) |
| **Physical Activity** | -.0248 (.0041, -6.09, <.001)* | -.0482 (.0236, -2.04, .041)* | – | – | .0012 (.0007, .0001 – .0027)* | .3% |
| **Weekend Screen Time (Reverse-Scored)** | -.0504 (.0059, -8.57, <.001)* | -.0131 (.0166, -.79, .43) | – | – | .0007 (.0010, -.0011 – .0027) | .2% (n.s.) |
| **All Mediators** | – | – | – | – | .0713 (.0062, .0595 – .0840)* | 21% |
| **CBCL Anxiety** | – | – | .3188 (.0082, 38.75, <.001)* | .2519 (.0086, 29.36, <.001)* | – | – |
| **Sleep Quality** | -.4628 (.0140, -33.09, <.001)* | -.1347 (.0067, -20.16, <.001)* | – | – | .0624 (.0051, .0528 – .0729)* | 19.6% |
| **Diet** | -.0380 (.0045, -8.49, <.001)* | -.0716 (.0211, -3.39, <.001)* | – | – | .0027 (.0009, .0011 – .0047)* | .8% |
| **Physical Activity** | -.0193 (.0040, -4.86, <.001)* | -.0534 (.0236, -2.26, <.024) | – | – | .0010 (.0005, .0002 – .0022)* | .3% |
| **Weekend Screen Time (Reverse-Scored)** | -.0253 (.0058, -4.40, <.001)* | -.0282 (.0166, -1.70, .089) | – | – | .0007 (.0005, -.0001 – .0020) | .2% (n.s.) |
| **All Mediators** | – | – | – | – | .0668 (.0052, .0573 – .0775)* | 21% |
| **PQ-BC Distress** | – | – | .0717 (.0063, 11.45, <.001)* | .0509 (.0059, 8.58, <.001)* | – | – |
| **Sleep Quality** | -.0952 (.0105, -9.11, <.001)* | -.1980 (.0066, -29.94, <.001)* | – | – | .0188 (.0027, .0138 – .0246)* | 26.2% |
| **Physical Activity** | -.0140 (.0028, -5.04, <.001)* | -.0736 (.0248, -2.97, .003)* | – | – | .0010 (.0004, .0003 – .0020)* | 1.4% |
| **Diet** | -.0050 (.0032, -1.59, .11) | -.1115 (.0222, -5.03, <.001)* | – | – | .0006 (.0004, -.0002 – .0015)* | .8% |
| **Weekend Screen Time (Reverse-Scored)** | -.0225 (.0040, -5.58, <.001)* | -.0178 (.0174, -1.02, .31) | – | – | .0004 (.0004, -.0004 – .0014) | .6% (n.s.) |
| **All Mediators** | – | – | – | – | .0208 (.0028, .0156 – .0268)* | 29% |

**Supplementary Table 6.** Results from mediation analyses in which Child Behavioral Checklist (CBCL) depression or anxiety t-scores or Prodromal Questionnaire-Brief Child (PQ-BC) version distress score at 1-year follow-up were used as predictors (independent variables), healthy lifestyle indicators (sleep quality (reverse score of the Sleep Disturbance Scale for Children, year 2), weekday screen time (reverse scored, year 2), physical activity (year 2), and Mediterranean diet (year 1)) as mediators, and social problems (CBCL Social Problems score) as the outcome (dependent variable). Covariates included age (year 2), sex, and site. %Mediation is calculated as Indirect Effect ÷ Total Effect × 100. **p* < .05. CBCL Total Problems was not used as a predictor as it includes CBCL social score when calculating its value. Mediators are listed in order of decreasing %mediation. Abbreviations: CI = Confidence Interval, n.s. = non-significant, SE = Standard Error.

| **Predictor** | **Predictor to Mediator B (SE, t, *p*)** | **Mediator to Outcome B (SE, t, *p*)** | **Total Effect, i.e., Predictor + Mediators to Outcome B (SE, t, *p*)** | **Direct Effect, i.e., Predictor to Outcome B (SE, t, *p*)** | **Indirect Effect, i.e., Mediation B (SE, Bootstrapped 95% CI)** | **%Mediation** |
| --- | --- | --- | --- | --- | --- | --- |
| **Mediator** |  |  |  |  |  |  |
| **CBCL Depression** | – | – | .3470 (.0084, 41.44, <.001)* | .2736 (.0092, 29.59, <.001)* | – | – |
| **Sleep Quality** | -.5845 (.0137, -42.67, <.001)* | -.1187 (.0069, -17.18, <.001)* | – | – | .0694 (.0063, .0576 – .0824)* | 20.0% |
| **Physical Activity** | -.0260 (.0041, -6.42, <.001)* | -.0797 (.0234, -3.40, <.001)* | – | – | .0021 (.0007, .0008 – .0036)* | .6% |
| **Diet** | -.0659 (.0045, -14.59, <.001)* | -.0269 (.0210, -1.28, .20) | – | – | .0018 (.0014, -.0010 – .0047) | .5% (n.s.) |
| **Weekday Screen Time (Reverse-Scored)** | -.0300 (.0059, -5.06, <.001)* | -.0070 (.0161, -.43, .67) | – | – | .0002 (.0005, -.0007 – .0014) | .1% (n.s.) |
| **All Mediators** | – | – | – | – | .0734 (.0065, .0617 – .0870)* | 21% |
| **CBCL Anxiety** | – | – | .3213 (.0082, 38.98, <.001)* | .2532 (.0086, 29.50, <.001)* | – | – |
| **Sleep Quality** | -.4595 (.0139, -33.09, <.001)* | -.1382 (.0067, -20.77, <.001)* | – | – | .0635 (.0051, .0540 – .0742)* | 19.8% |
| **Diet** | -.0379 (.0044, -8.53, <.001)* | -.0687 (.0209, -3.29, .001)* | – | – | .0026 (.0009, .0010 – .0045)* | .8% |
| **Physical Activity** | -.0210 (.0040, -5.31, <.001)* | -.0849 (.0234, -3.62, <.001) | – | – | .0018 (.0006, .0008 – .0033)* | .6% |
| **Weekday Screen Time (Reverse-Scored)** | -.0138 (.0058, -2.39, .017)* | -.0174 (.0161, -1.08, .28) | – | – | .0002 (.0003, -.0001 – .0012) | .1% (n.s.) |
| **All Mediators** | – | – | – | – | .0681 (.0052, .0584 – .0786)* | 21% |
| **PQ-BC Distress** | – | – | .0808 (.0061, 13.15, <.001)* | .0597 (.0058, 10.26, <.001)* | – | – |
| **Sleep Quality** | -.0947 (.0102, -9.32, <.001)* | -.1997 (.0066, -30.36, <.001)* | – | – | .0189 (.0027, .0141 – .0247)* | 23.4% |
| **Physical Activity** | -.0174 (.0027, -6.39, <.001)* | -.1020 (.0246, -4.15, <.001)* | – | – | .0018 (.0005, .0009 – .0030)* | 2.2% |
| **Diet** | -.0029 (.0031, -.93, .35) | -.1093 (.0219, -5.00, <.001)* | – | – | .0003 (.0004, -.0004 – .0012)* | .4% (n.s.) |
| **Weekday Screen Time (Reverse-Scored)** | -.0194 (.0040, -4.87, <.001)* | -.0073 (.0169, -.43, .66) | – | – | .0001 (.0004, -.0005 – .0009) | .1% (n.s.) |
| **All Mediators** | – | – | – | – | .0211 (.0028, .0161 – .0270)* | 26% |

**Supplementary Table 7.** Effects of environmental moderators on the relationships between lifestyle mediators and academic functioning (school grades between follow-up years 2-3). Notably, ΔR^2^ represents the change in total variance explained due to the moderator, and moderators associated with small ΔR^2^ values can still show robust, significant effects on %mediation. F (*p*) values are for the highest order unconditional interactions. Only healthy lifestyle factors that showed significant mediation were examined for moderation effects. **p* < .05 for the moderated mediation effect.

| **Predictor** | **Moderator** | **ΔR^2^** | **F (*p*)** | **Index of Moderated Mediation** | | |
| --- | --- | --- | --- | --- | --- | --- |
| **Mediator** |  |  |  | **Index** | **SE** | **95% CI** |
| **CBCL Depression** | | | | | | |
| **Sleep Quality** | Neighborhood Safety | .0002 | 1.23 (.27) | -.0022 | .0024 | -.0067 – .0028 |
| **Weekend Screen Time (Reverse-Scored)** |  | .0000 | .33 (.57) | -.0003 | .0006 | -.0014 – .0008 |
| **Physical Activity** |  | .0005 | 4.03 (.045) | .0007 | .0004 | .0000 – .0015 |
| **Diet** |  | – | – | – | – | – |
| **Sleep Quality** | Family Conflict | .0001 | .43 (.51) | -.0006 | .0011 | -.0027 – .0015 |
| **Weekend Screen Time (Reverse-Scored)** |  | .0003 | 2.50 (.11) | -.0004 | .0003 | -.0010 – .0002 |
| **Physical Activity** |  | .0000 | .15 (.70) | -.0001 | .0002 | -.0004 – .0003 |
| **Diet** |  | – | – | – | – | – |
| **Sleep Quality** | School Environment | .0004 | 3.00 (.083) | -.0012 | .0008 | -.0029 – .0004 |
| **Weekend Screen Time (Reverse-Scored)** |  | .0002 | 1.17 (.28) | .0002 | .0002 | -.0002 – .0006 |
| **Physical Activity** |  | .0000 | .08 (.78) | .0000 | .0001 | -.0002 – .0003 |
| **Diet** |  | – | – | – | – | – |
| **Sleep Quality** | Family Financial Adversity | .0010 | 7.52 (.006) | .0044 | .0019 | .0005 – .0081* |
| **Weekend Screen Time (Reverse-Scored)** |  | .0016 | 12.12 (<.001) | .0013 | .0005 | .0005 - .0023* |
| **Physical Activity** |  | .0000 | .13 (.72) | -.0001 | .0003 | -.0008 – .0006 |
| **Diet** |  | – | – | – | – | – |
| **CBCL Anxiety** | | | | | | |
| **Sleep Quality** | Neighborhood Safety | .0002 | 1.60 (.21) | -.0020 | .0019 | -.0057 – .0017 |
| **Weekend Screen Time (Reverse-Scored)** |  | .0000 | .14 (.71) | -.0001 | .0003 | -.0007 – .0004 |
| **Physical Activity** |  | .0005 | 3.64 (.056) | .0005 | .0003 | .0000 – .0013 |
| **Diet** |  | .0000 | .00 (.99) | .0000 | .0005 | -.0009 – .0009 |
| **Sleep Quality** | Family Conflict | .0001 | .58 (.45) | -.0006 | .0009 | -.0022 – .0012 |
| **Weekend Screen Time (Reverse-Scored)** |  | .0004 | 2.56 (.11) | -.0002 | .0001 | -.0005 – .0001 |
| **Physical Activity** |  | .0000 | .12 (.73) | .0000 | .0001 | -.0003 – .0002 |
| **Diet** |  | .0000 | .06 (.80) | -.0001 | .0002 | -.0005 – .0004 |
| **Sleep Quality** | School Environment | .0004 | 3.02 (.082) | -.0009 | .0007 | -.0022 – .0003 |
| **Weekend Screen Time (Reverse-Scored)** |  | .0002 | 1.58 (.21) | .0001 | .0001 | -.0001 – .0003 |
| **Physical Activity** |  | .0000 | .13 (.72) | .0000 | .0001 | -.0002 – .0003 |
| **Diet** |  | .0003 | 1.89 (.17) | .0002 | .0002 | -.0001 – .0006 |
| **Sleep Quality** | Family Financial Adversity | .0011 | 8.10 (.004) | .0036 | .0015 | .0005 – .0066* |
| **Weekend Screen Time (Reverse-Scored)** |  | .0015 | 10.72 (.001) | .0006 | .0003 | .0002 – .0012* |
| **Physical Activity** |  | .0000 | .14 (.71) | -.0001 | .0003 | -.0007 – .0004 |
| **Diet** |  | .0005 | 3.38 (.066) | -.0007 | .0004 | -.0015 – .0002 |
| **PQ-BC Distress** | | | | | | |
| **Sleep Quality** | Neighborhood Safety | .0002 | 1.36 (.24) | -.0004 | .0004 | -.0012 – .0004 |
| **Weekend Screen Time (Reverse-Scored)** |  | .0000 | .17 (.68) | -.0001 | .0003 | -.0008 – .0005 |
| **Physical Activity** |  | .0006 | 4.34 (.037) | .0005 | .0003 | .0000 – .0011 |
| **Diet** |  | – | – | – | – | – |
| **Sleep Quality** | Family Conflict | .0001 | .71 (.41) | -.0001 | .0002 | -.0005 – .0002 |
| **Weekend Screen Time (Reverse-Scored)** |  | .0005 | 3.79 (.052) | -.0002 | .0002 | -.0006 – .0000 |
| **Physical Activity** |  | .0000 | .13 (.72) | .0000 | .0001 | -.0003 – .0002 |
| **Diet** |  | – | – | – | – | – |
| **Sleep Quality** | School Environment | .0003 | 2.35 (.13) | -.0002 | .0001 | -.0004 – .0001 |
| **Weekend Screen Time (Reverse-Scored)** |  | .0002 | 1.15 (.28) | .0001 | .0001 | -.0001 – .0003 |
| **Physical Activity** |  | .0000 | .07 (.80) | .0000 | .0001 | -.0002 – .0002 |
| **Diet** |  | – | – | – | – | – |
| **Sleep Quality** | Family Financial Adversity | .0011 | 8.47 (.004) | .0007 | .0003 | .0001 – .0014* |
| **Weekend Screen Time (Reverse-Scored)** |  | .0012 | 9.09 (.003) | .0006 | .0003 | .0002 – .0012* |
| **Physical Activity** |  | .0000 | .09 (.76) | -.0001 | .0002 | -.0005 – .0004 |
| **Diet** |  | – | – | – | – | – |
| **CBCL Total Problems** | | | | | | |
| **Weekend Screen Time (Reverse-Scored)** | Neighborhood Safety | .0001 | .03 (.86) | -.0002 | .0004 | -.0009 – .0005 |
| **Sleep Quality** |  | – | – | – | – | – |
| **Physical Activity** |  | .0005 | 3.75 (.053) | .0003 | .0002 | .0000 – .0008 |
| **Diet** |  | – | – | – | – | – |
| **Weekend Screen Time (Reverse-Scored)** | Family Conflict | .0004 | 3.13 (.077) | -.0003 | .0002 | -.0006 – .0001 |
| **Sleep Quality** |  | – | – | – | – | – |
| **Physical Activity** |  | .0000 | .13 (.72) | .0000 | .0001 | -.0002 – .0002 |
| **Diet** |  | – | – | – | – | – |
| **Weekend Screen Time (Reverse-Scored)** | School Environment | .0002 | 1.87 (.17) | .0001 | .0001 | -.0001 – .0004 |
| **Sleep Quality** |  | – | – | – | – | – |
| **Physical Activity** |  | .0000 | .10 (.76) | .0000 | .0001 | -.0001 – .0002 |
| **Diet** |  | – | – | – | – | – |
| **Weekend Screen Time (Reverse-Scored)** | Family Financial Adversity | .0014 | 10.89 (.001) | .0007 | .0003 | .0003 – .0013* |
| **Sleep Quality** |  | – | – | – | – | – |
| **Physical Activity** |  | .0001 | .84 (.36) | -.0001 | .0002 | -.0005 – .0002 |
| **Diet** |  | – | – | – | – | – |

**Supplementary Table 8.** Effects of environmental moderators on the relationships between lifestyle mediators and social problems (CBCL Social Problems score at year 3). Notably, ΔR^2^ represents the change in total variance explained due to the moderator, and moderators associated with small ΔR^2^ values can still show robust, significant effects on %mediation. F (*p*) values are for the highest order unconditional interactions. Only healthy lifestyle factors that showed significant mediation were examined for moderation effects. **p* < .05 for the moderated mediation effect.

| **Predictor** | **Moderator** | **ΔR^2^** | **F (*p*)** | **Index of Moderated Mediation** | | |
| --- | --- | --- | --- | --- | --- | --- |
| **Mediator** |  |  |  | **Index** | **SE** | **95% CI** |
| **CBCL Depression** | | | | | | |
| **Sleep Quality** | Neighborhood Safety | .0001 | .63 (.43) | .0028 | .0051 | -.0075 – .0126 |
| **Weekend Screen Time (Reverse-Scored)** |  | – | – | – | – | – |
| **Physical Activity** |  | .0002 | 2.02 (.16) | -.0009 | .0007 | -.0025 – .0004 |
| **Diet** |  | – | – | – | – | – |
| **Sleep Quality** | Family Conflict | .0019 | 18.68 (<.001) | .0072 | .0022 | .0027 – .0115* |
| **Weekend Screen Time (Reverse-Scored)** |  | – | – | – | – | – |
| **Physical Activity** |  | .0000 | .43 (.51) | .0002 | .0004 | -.0005 – .0009 |
| **Diet** |  | – | – | – | – | – |
| **Sleep Quality** | School Environment | .0011 | 10.42 (.001) | -.0040 | .0017 | -.0072 – -.0006* |
| **Weekend Screen Time (Reverse-Scored)** |  | – | – | – | – | – |
| **Physical Activity** |  | .0000 | .01 (.91) | .0000 | .0002 | -.0004 – .0005 |
| **Diet** |  | – | – | – | – | – |
| **Sleep Quality** | Family Financial Adversity | .0019 | 18.37 (<.001) | .0117 | .0051 | .0020 – .0219* |
| **Weekend Screen Time (Reverse-Scored)** |  | – | – | – | – | – |
| **Physical Activity** |  | .0001 | 1.19 (.28) | -.0006 | .0008 | -.0023 – .0009 |
| **Diet** |  | – | – | – | – | – |
| **CBCL Anxiety** | | | | | | |
| **Sleep Quality** | Neighborhood Safety | .0001 | .98 (.32) | .0028 | .0041 | -.0054 – .0108 |
| **Weekend Screen Time (Reverse-Scored)** |  | .0001 | .57 (.45) | -.0003 | .0006 | -.0016 – .0007 |
| **Physical Activity** |  | .0001 | 1.44 (.23) | -.0006 | .0006 | -.0020 – .0004 |
| **Diet** |  | .0004 | 4.16 (.041) | -.0016 | .0009 | -.0035 – .0001 |
| **Sleep Quality** | Family Conflict | .0017 | 16.59 (<.001) | .0053 | .0017 | .0022 – .0087* |
| **Weekend Screen Time (Reverse-Scored)** |  | .0002 | 1.61 (.20) | .0003 | .0003 | -.0002 – .0009 |
| **Physical Activity** |  | .0000 | .34 (.56) | .0001 | .0003 | -.0004 – .0008 |
| **Diet** |  | .0004 | 3.91 (.048) | .0008 | .0005 | -.0001 – .0017 |
| **Sleep Quality** | School Environment | .0008 | 8.16 (.004) | -.0028 | .0013 | -.0054 – -.0001* |
| **Weekend Screen Time (Reverse-Scored)** |  | .0007 | 7.02 (.008) | .0004 | .0002 | .0001 – .0009* |
| **Physical Activity** |  | .0000 | .00 (.96) | .0000 | .0002 | -.0004 – .0004 |
| **Diet** |  | .0001 | .62 (.43) | -.0002 | .0003 | -.0009 – .0003 |
| **Sleep Quality** | Family Financial Adversity | .0020 | 19.39 (<.001) | .0095 | .0040 | .0021 – .0175* |
| **Weekend Screen Time (Reverse-Scored)** |  | .0001 | .88 (.35) | .0003 | .0005 | -.0005 – .0016 |
| **Physical Activity** |  | .0001 | 1.10 (.29) | -.0005 | .0006 | -.0018 – .0007 |
| **Diet** |  | .0001 | .50 (.48) | .0005 | .0009 | -.0014 – .0023 |
| **PQ-BC Distress** | | | | | | |
| **Sleep Quality** | Neighborhood Safety | .0002 | 1.67 (.20) | .0008 | .0009 | -.0009 – .0028 |
| **Weekend Screen Time (Reverse-Scored)** |  | – | – | – | – | – |
| **Physical Activity** |  | .0003 | 2.24 (.13) | -.0007 | .0005 | -.0018 – .0003 |
| **Diet** |  | – | – | – | – | – |
| **Sleep Quality** | Family Conflict | .0025 | 22.57 (<.001) | .0013 | .0004 | .0006 – .0023* |
| **Weekend Screen Time (Reverse-Scored)** |  | – | – | – | – | – |
| **Physical Activity** |  | .0000 | .32 (.57) | .0001 | .0003 | -.0003 – .0007 |
| **Diet** |  | – | – | – | – | – |
| **Sleep Quality** | School Environment | .0011 | 9.31 (.002) | -.0006 | .0003 | -.0012 – .0001* |
| **Weekend Screen Time (Reverse-Scored)** |  | – | – | – | – | – |
| **Physical Activity** |  | .0000 | .00 (.99) | .0000 | .0002 | -.0003 – .0003 |
| **Diet** |  | – | – | – | – | – |
| **Sleep Quality** | Family Financial Adversity | .0014 | 12.04 (<.001) | .0016 | .0009 | .0001 – .0037* |
| **Weekend Screen Time (Reverse-Scored)** |  | – | – | – | – | – |
| **Physical Activity** |  | .0002 | 1.29 (.26) | -.0004 | .0005 | -.0016 – .0005 |
| **Diet** |  | – | – | – | – | – |

**Supplementary Figure 1.** Diagram illustrating moderation analysis procedure. The effect of each individual moderator (e.g., family financial adversity) was tested on the relationships between all mediators (e.g., sleep quality) and each outcome (e.g., academic functioning). In this fashion, moderated mediation models examined the degree to which each moderator affected the mediation of associations between mental health with outcome through lifestyle factors.

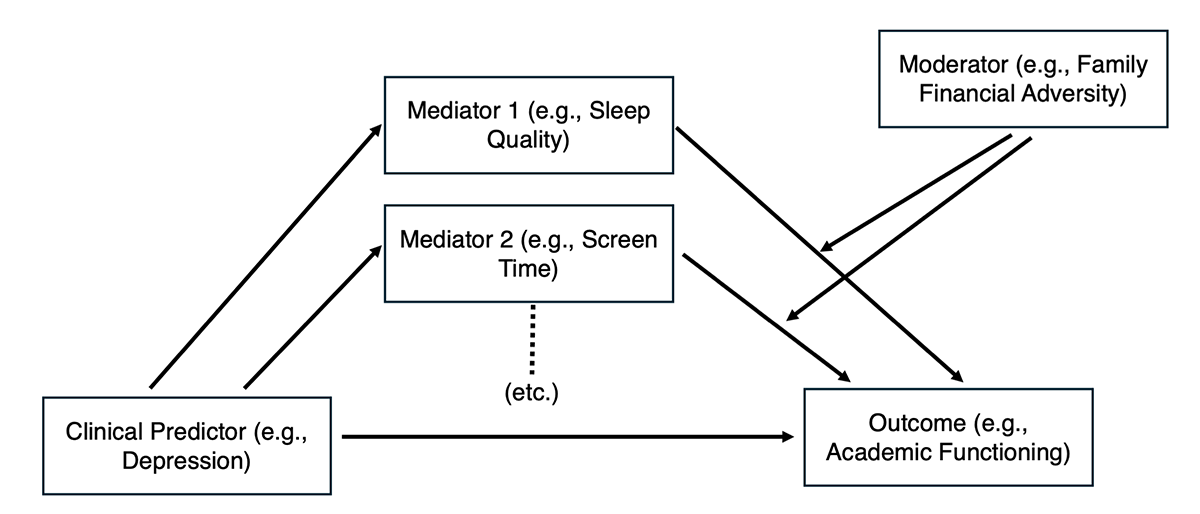

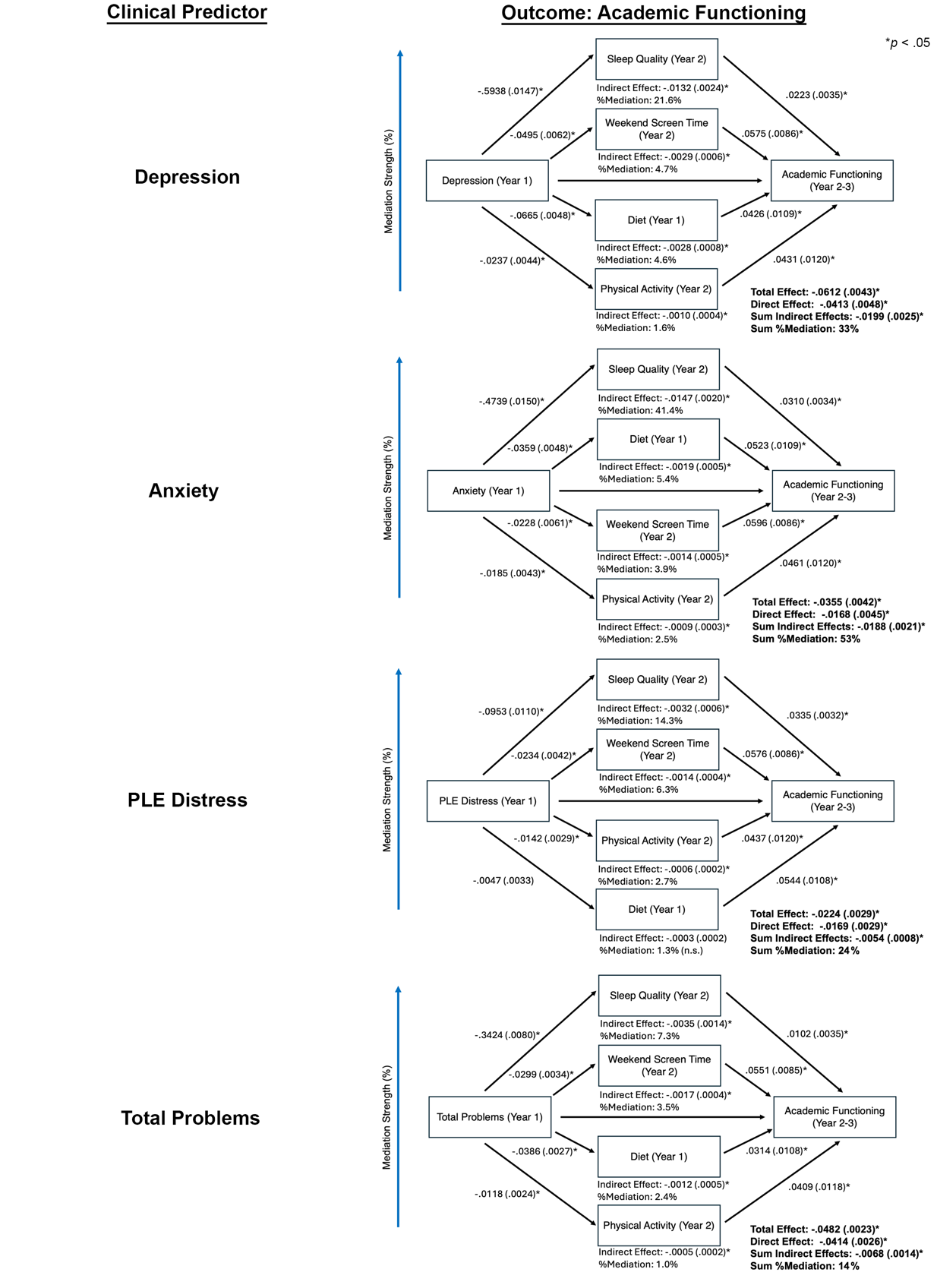
**Supplementary Figure 2.** Results of mediation models predicting academic functioning (school grades between follow-up years 2-3) while including age (year 2), sex, site, and National Institutes of Health Cognition Toolbox Total Composite raw score as covariates. More detailed results are provided in Supplementary Table 2. PLE = Psychoticlike Experience.

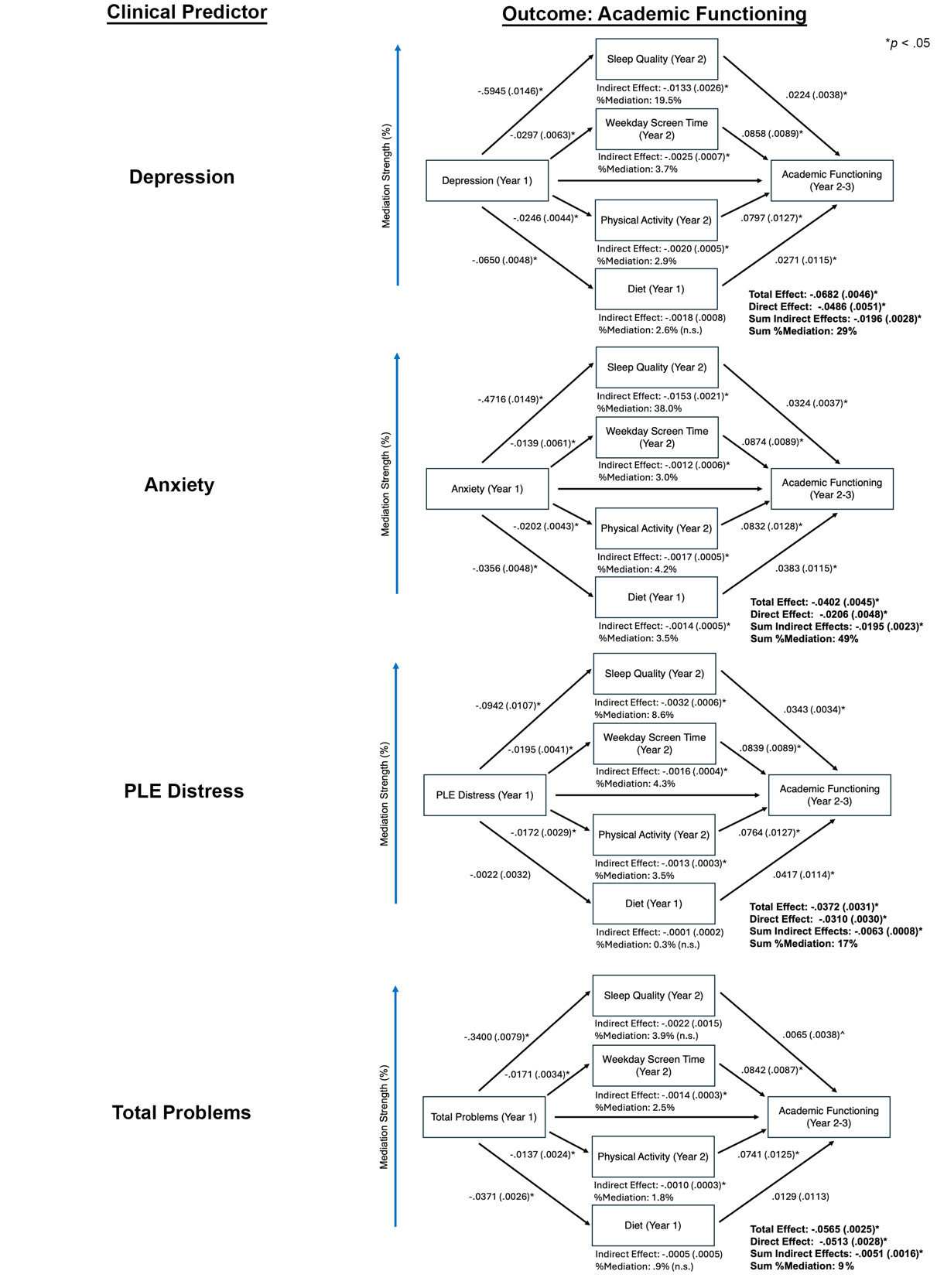
**Supplementary Figure 3.** Results of mediation models predicting academic functioning (school grades between follow-up years 2-3) while including age (year 2), sex, and site as covariates and weekday (as opposed to weekend) screen time as a mediator. More detailed results are provided in Supplementary Table 3. PLE = Psychoticlike Experience.

**Supplementary Figure 4.** Results of mediation models predicting social problems (Child Behavior Checklist Social Problems score at year 3) while including age (year 2), sex, and site, and National Institutes of Health Cognition Toolbox Total Composite raw score as covariates. More detailed results are provided in Supplementary Table 5. PLE = Psychoticlike Experience.

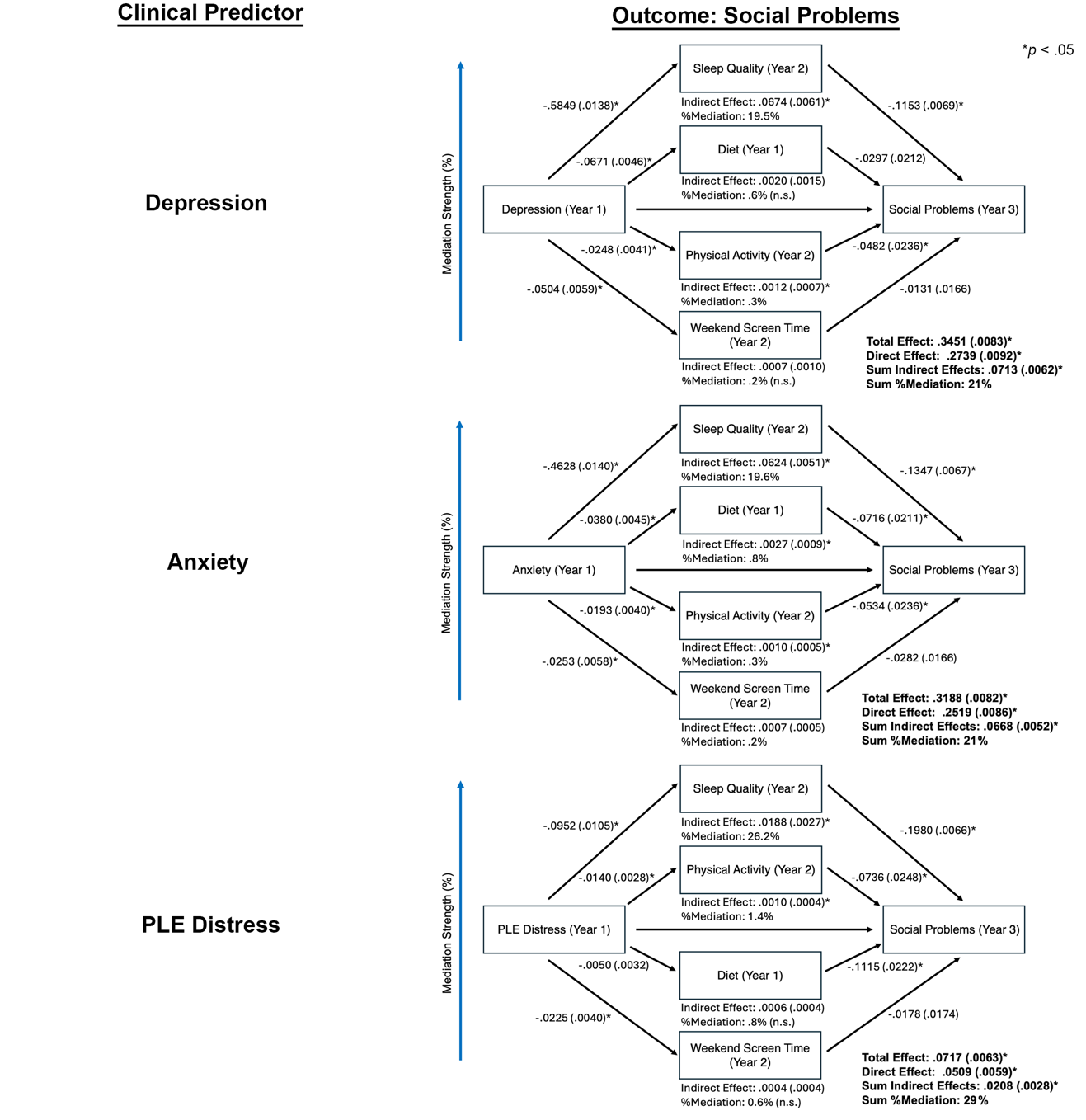

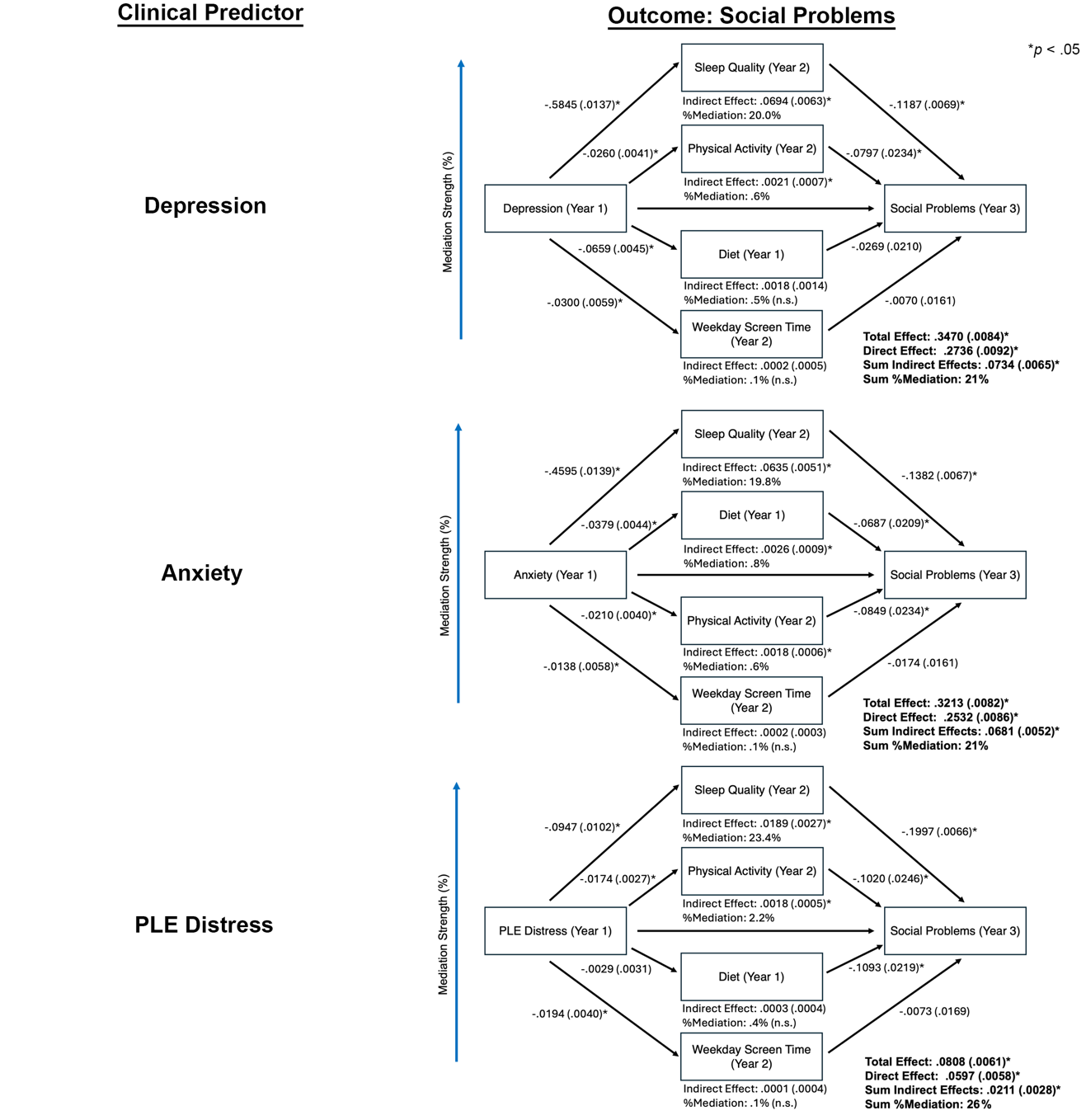
**Supplementary Figure 5.** Results of mediation models predicting social problems (Child Behavior Checklist Social Problems score at year 3) while including age (year 2), sex, and site as covariates and weekday (as opposed to weekend) screen time as a mediator. More detailed results are provided in Supplementary Table 6. PLE = Psychoticlike Experience.

**References**

1. Nagata JM, Bashir A, Weinstein S, Al-Shoaibi AAA, Shao IY, Ganson KT, Testa A, Garber AK. Social epidemiology of the Mediterranean-dietary approaches to stop hypertension intervention for neurodegenerative delay (MIND) diet among early adolescents: the Adolescent Brain Cognitive Development Study. Pediatr Res. 2024;96:230-236.

2. Livingstone MB, Robson PJ, Wallace JM. Issues in dietary intake assessment of children and adolescents. Br J Nutr. 2004;92 Suppl 2:S213-222.

3. Jones L, Ness A, Emmett P. Misreporting of Energy Intake From Food Records Completed by Adolescents: Associations With Sex, Body Image, Nutrient, and Food Group Intake. Front Nutr. 2021;8:749007.

4. Wallace A, Kirkpatrick SI, Darlington G, Haines J. Accuracy of Parental Reporting of Preschoolers' Dietary Intake Using an Online Self-Administered 24-h Recall. Nutrients. 2018;10.

5. Bruni O, Ottaviano S, Guidetti V, Romoli M, Innocenzi M, Cortesi F, Giannotti F. The Sleep Disturbance Scale for Children (SDSC). Construction and validation of an instrument to evaluate sleep disturbances in childhood and adolescence. J Sleep Res. 1996;5:251-261.

6. Echeverria SE, Diez-Roux AV, Link BG. Reliability of self-reported neighborhood characteristics. J Urban Health. 2004;81:682-701.

7. Moos RH: Family Environment Scale Manual: Development, Applications, Research, Consult. Psychol. Press; 1994.

8. Zucker RA, Gonzalez R, Feldstein Ewing SW, Paulus MP, Arroyo J, Fuligni A, Morris AS, Sanchez M, Wills T. Assessment of culture and environment in the Adolescent Brain and Cognitive Development Study: Rationale, description of measures, and early data. Dev Cogn Neurosci. 2018;32:107-120.

9. Karcher NR, Loewy RL, Savill M, Avenevoli S, Huber RS, Simon TJ, Leckliter IN, Sher KJ, Barch DM. Replication of Associations With Psychotic-Like Experiences in Middle Childhood From the Adolescent Brain Cognitive Development (ABCD) Study. Schizophr Bull Open. 2020;1:sgaa009.
